# Dominant truncating variants in *KAT6A* cause two neurodevelopmental disorders with opposite gene regulatory and metabolic changes

**DOI:** 10.64898/2026.08.11.26358095

**Authors:** Aileen A. Nava, Yaneth Perez-Rodriguez, Tzung-Chien Hsieh, Alexander S. Byrne, Abigail S. Krall, Jerome Freudenberg, Niloufar Mansooralavi, Vijaya Pandey, Linsey Stiles, Cristiane Benincá, Jing-Mei Li, Sanaa Choufani, Meghna Singh, Shahida Moosa, Irene Valenzuela, Eduardo F. Tizzano, Amélie Piton, Didier Lacombe, Laurence Perrin, Jesus Marquez, Juan Dario Ortigoza-Escobar, Sophia Ahmadyar, Harold Pimentel, James A. Wohlschlegel, Luis de la Torre-Ubieta, Heather R. Christofk, Rosanna Weksberg, William E. Lowry, Valerie A. Arboleda

## Abstract

GRAPHICAL ABSTRACT

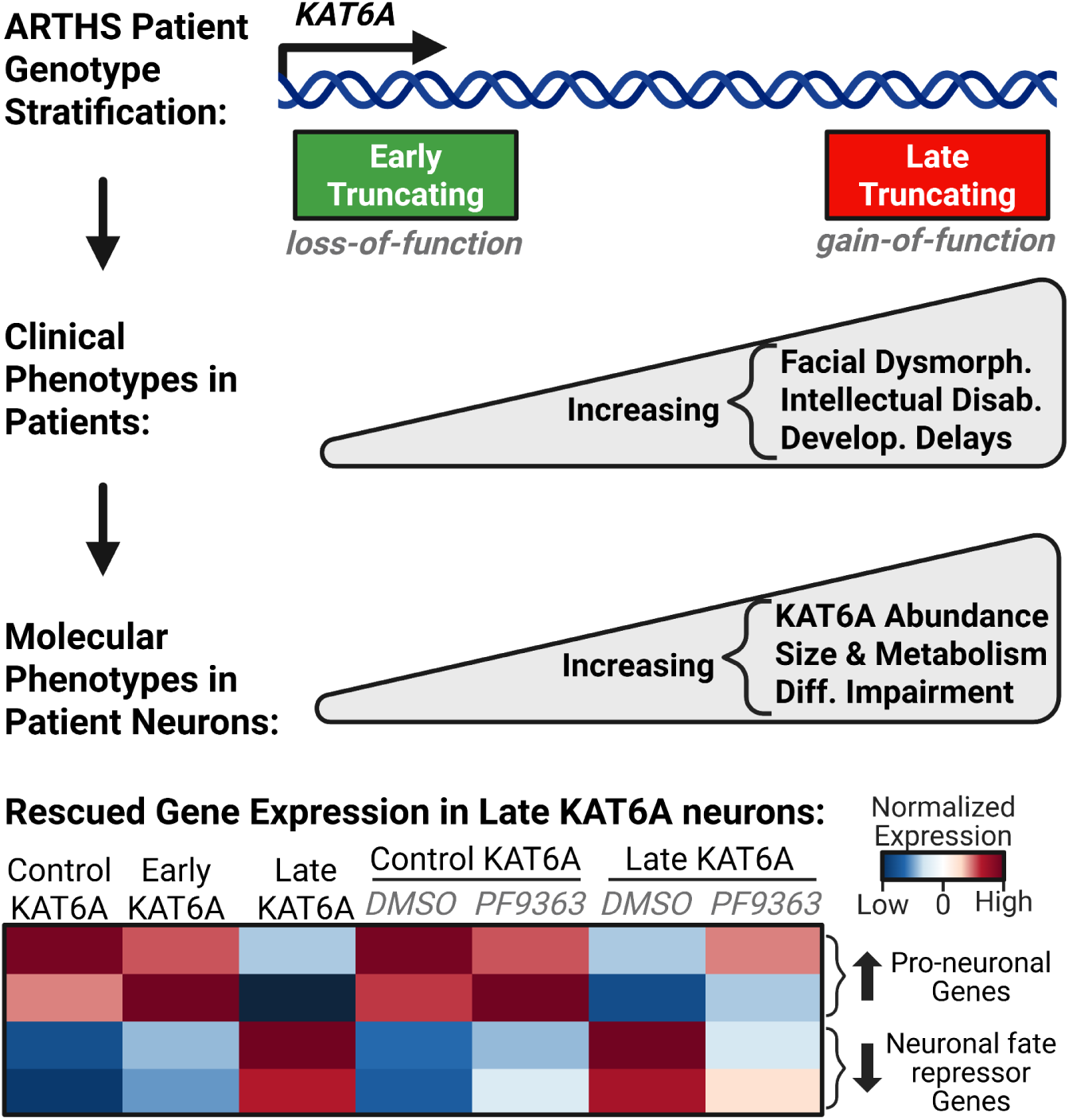

**KEY POINTS:**

- Truncating variant location within *KAT6A* determines outcome: nonsense-mediated decay either silences early-truncating variants or spares late-truncating variants, producing loss- or gain-of-function and two distinct neurodevelopmental disorders.
- Early- and late-truncating *KAT6A* variants affect gene regulation in opposite directions centered on a *KAT6A*-*FOS*-*RBFOX*-*PTBP* axis, stalling development at distinct stages.
- Early- and late-truncating *KAT6A* variants drive inverse metabolic states in neurons via a multi-tiered mechanism, unveiling *KAT6A*’s complex role in human metabolism.
- Pairing clinical patient assessment with multi-omic profiling of patient-derived stem cells is a powerful strategy for cracking pathogenic mechanisms in rare Mendelian disease.
- Chemical inhibitors targeting epigenes can be used as disease-modifying strategies for Chromatinopathies, but only if their underlying pathogenic mechanism is known.

Arboleda-Tham Syndrome (ARTHS), caused by truncating variants in KAT6A, is currently diagnosed as a single neurodevelopmental syndrome with variable severity of intellectual disability and multi-system findings. Here, we reveal that this clinical stratification reflects fundamentally distinct molecular mechanisms driven by variant position in the gene. Using patient-derived iPSCs and multi-omics profiling, we demonstrate that early-truncating variants (exons 1-15) cause loss-of-function via nonsense-mediated decay (NMD), while late-truncating variants (exons 16-17) that escape NMD cause gain-of-function effects. These opposite mechanisms are reflected in distinctive facial gestalt features and DNA-methylation episignatures and invert the direction of change across neuronal gene regulation, metabolism, and mitochondrial physiology. This mechanistic distinction enables precision therapeutics: late-truncating variants are amenable to *KAT6A* inhibition, while early-truncating variants require loss-of-function rescue. Variant-level stratification is therefore essential: mechanistic understanding—not gene-level diagnosis alone—is prerequisite for developing rational therapeutic strategies in rare Mendelian disease.

## INTRODUCTION

Arboleda-Tham Syndrome (ARTHS, MIM#616268) is a rare neurodevelopmental disorder caused by heterozygous *Lysine (K) Acetyltransferase 6A* (*KAT6A;* a.k.a. *MYST3*, *MOZ*) variants^1–3^. Pathogenic germline *KAT6A* variants are predominantly nonsense or frameshift mutations creating premature termination codons (PTCs). Prior work demonstrated a genotype-phenotype association in ARTHS: early truncating variants (exons 1–15, affecting residues 1–1013) associate with mild intellectual disability and subtle dysmorphic features, while late truncating variants (exons 16–17, affecting residues 1014–2004) predict severe multisystem neurodevelopmental disease^4^. While this observation has been independently validated by other groups^5,6^, the clinical syndrome remains grouped as a single uniform entity, despite the clinical heterogeneity. The molecular mechanisms explaining why variant location drives such divergent outcomes remain unknown. Understanding these mechanisms is essential for developing rational, variant-specific therapeutics and is the central motivation for the current study.

We previously postulated that nonsense-mediated decay (NMD) might explain the divergent outcomes. NMD is a quality-control mechanism that degrades mRNAs containing premature termination codons (PTCs). Critically, the position of the PTC relative to exon-exon junctions determines NMD sensitivity: PTCs falling ≤50–55 nucleotides upstream of the terminal exon-exon junction, or PTCs occurring within the final exon, escape NMD ^7^.

Accordingly, across multiple Mendelian syndromes caused by *SOX10*, *MPZ*, and *MN1*, 5’-localized PTCs trigger NMD-mediated haploinsufficiency, while 3’-localized PTCs escape NMD and cause dominant-negative or gain-of-function effects that produce distinct clinical syndromes^8–12^. These precedents established that variant location can fundamentally alter disease mechanism and clinical outcome, raising the possibility that *KAT6A* follows a similar paradigm.

To investigate these mechanisms, we established patient-derived iPSC lines spanning early- and late-truncating variants and differentiated them through neural progenitor cells to neurons over 70 days in vitro. We applied multi-omics profiling — spanning chromatin accessibility, transcriptome, proteome, and metabolome — across differentiation stages to trace how variant-location-dependent differences in KAT6A abundance propagate through neurodevelopmental gene regulatory networks.

Our analysis revealed that early- and late-truncating *KAT6A* variants cause loss-of-function and gain-of-function, respectively, through differential NMD-mediated *KAT6A* abundance. These opposite molecular phenotypes manifest across gene regulation, neuronal morphology, and metabolic function during *in vitro* neurodevelopment. We further demonstrate that these mechanistic differences are stratified by accessible clinical biomarkers — facial gestalt analysis and blood-derived DNA methylation episignatures — and are pharmacologically rescuable selectively in late-truncating neurons using a A/B Histone acetyltransferase (HAT) KAT6 inhibitor, PF-9363^13–15^. Our findings establish that variant-location stratification in *KAT6A* reflects fundamentally distinct mechanistic pathways that demand precision medicine approaches—informing not only diagnosis but therapeutic strategy selection.

## RESULTS

### Facial phenotypes and episignatures stratify early- and late-truncating KAT6A variants

To determine whether variant position stratifies ARTHS clinical phenotypes, we applied GestaltMatcher facial analysis and DNA methylation episignature profiling to two independently collected patient cohorts — one providing facial photographs (n = 63) and one providing whole blood for DNA methylation profiling. (**Fig. 1A., Extended Data Fig. 1A-B**).

**Fig. 1:**
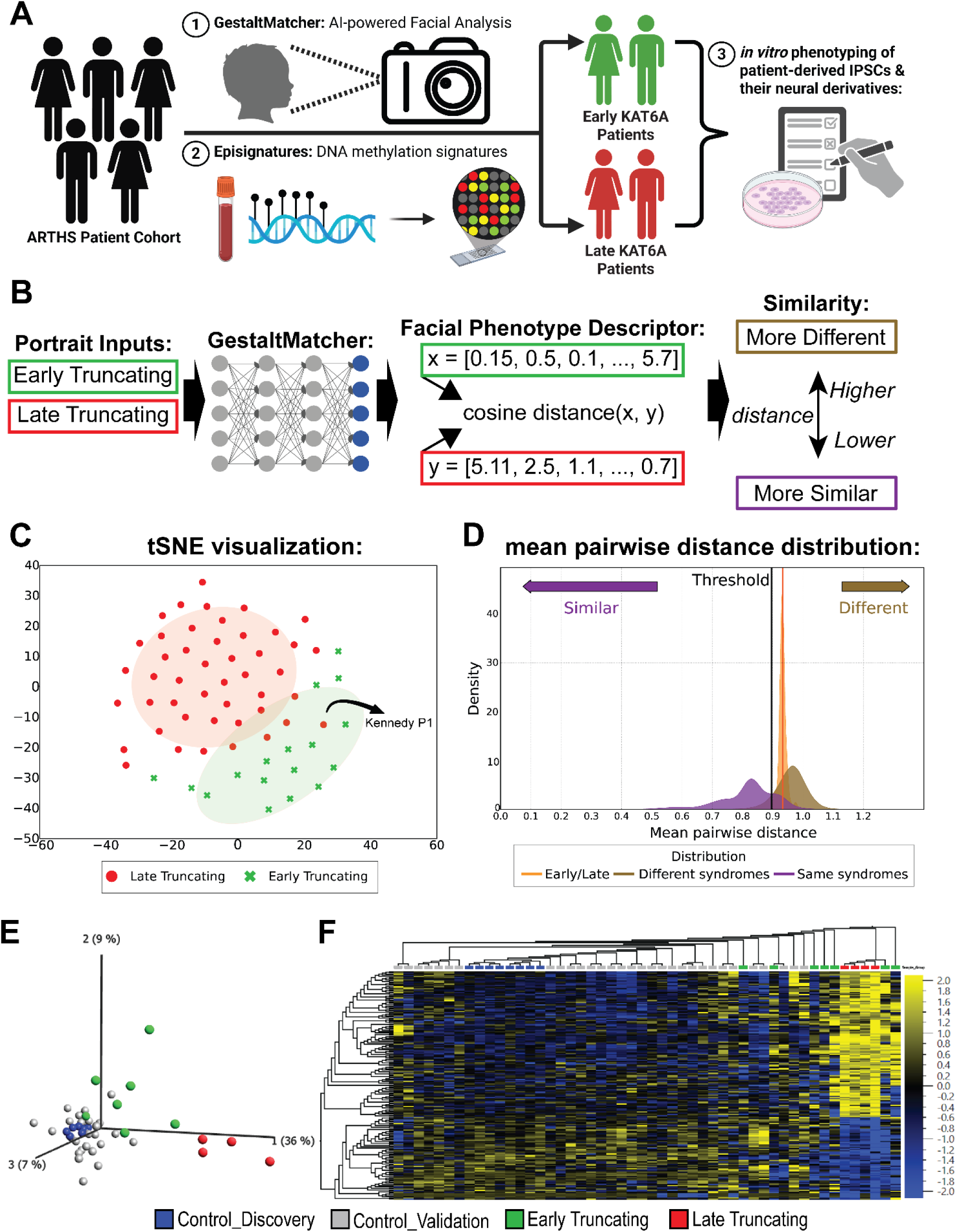
Early- and late-truncating KAT6A variants exhibit distinct facial phenotypes and DNA methylation signatures. **A**, Study overview from clinical assessments (GestaltMatcher facial analysis and episignature profiling) to comprehensive multi-omic profiling of patient-derived iPSCs and neural derivatives across early-truncating (green) and late-truncating (red) *KAT6A* variants. **B**, Schematic of GestaltMatcher approach: patient facial photographs are encoded into high-dimensional feature vectors (examples x and y) with pairwise cosine distances quantifying facial similarity (lower = more similar; higher = more different). **C**, t-SNE projection of facial phenotype embeddings from 63 ARTHS patients (17 early-truncating in green, 46 late-truncating in red), revealing distinct clustering by variant class. Shaded ellipses denote 95% confidence regions for each group. The most 3’ truncating *KAT6A* variant (Kennedy P1, annotated) clusters with early-truncating cases, suggesting there may be a second *KAT6A* boundary. Individual points = individual ARTHS patients. **D**, Distribution of mean pairwise distances for three comparisons: early vs. late truncating in ARTHS (orange), within-syndrome pairs (purple), and different-syndrome reference pairs (brown). Early–late mean distance of 0.933 exceeds the dissimilarity threshold (c = 0.896) in all 100 samples (range 0.915 – 0.969), with positive predictive value of 0.760. Linear SVM classification achieves 87.3% accuracy, 83.9% balanced accuracy, and 0.935 AUC. **E**, Principal component analysis (PCA) of 198 CpG sites identified in the late-truncating discovery cohort (4 late-truncating patients and 8 age/sex-matched discovery controls) using linear regression (FDR-corrected p < 0.05; |Δβ| > 0.10), visualized across all DNAm samples: 4 late-truncating patients (red), 8 discovery controls (blue), 7 early-truncating patients (green), and 31 validation controls (gray). Late-truncating profiles cluster distinctly from all control and early-truncating profiles. Individual points = individual ARTHS patients or controls. **F**, Unsupervised hierarchical clustering of the same 198 discovery CpG sites visualized across all DNAm samples (4 late-truncating patients, 7 early-truncating patients, 8 discovery controls, 31 validation controls). Of the 198 sites, 124 are hypermethylated and 74 are hypomethylated in late-truncating patients relative to controls. Individual columns = individual ARTHS patients or controls.

Using GestaltMatcher,^16^ which encodes facial portraits as high-dimensional phenotype descriptors mapped to a common Clinical Face Phenotype Space (**Fig. 1B)**, ^17^ we projected facial images of 63 ARTHS patients (17 early-truncating, 46 late-truncating)—revealing two visually-separated clusters driven by truncating variant class in t-SNE space (**Fig. 1C**). Pairwise cosine distance confirmed inter-group separation, and linear SVM classification achieved 87.3% accuracy (AUC = 0.935) (**Fig. 1D**). One late-truncating case, Kennedy P1 carrying *KAT6A* p.N1836Lfs*15 and only ∼170 amino acids from the end of the protein, projected closer to the early-truncating cluster. Kennedy P1 showed the most negative SVM decision score (−0.876; **Extended Data Fig. 1C**), suggesting a second *KAT6A* boundary after which cases have a milder phenotype, analogous to that seen in *KAT6B* disorders^18,19^.

We developed a variant class DNA methylation (DNAm) episignature from whole blood from 4 late-truncating patients and 8 matched controls. Linear regression modeling identified 198 differentially methylated CpG sites (FDR-corrected p < 0.05; |Δβ| > 0.10) of which 124 CpG sites were hypermethylated and 74 were hypomethylated.

To assess episignature specificity, we profiled genome-wide DNAm in an independent validation cohort of 31 controls and 7 early-truncating patients. We plotted the signature CpG sites for all DNAm samples using both principal component analysis (PCA) and hierarchical clustering (**Fig. 1E-F**). All 4 late-truncating patients clustered distinctly from control profiles. In contrast, early-truncating profiles were less clearly separated from controls, with some samples clustering near controls and others at intermediate positions between the late-truncating and control groups. Variant-class-specific chromatin states are therefore detectable in accessible clinical biospecimens; greater heterogeneity among early-truncating profiles likely reflects retained *KAT6A* function and variable epigenetic penetrance.

### Establishment of an in vitro model of neurodevelopment for ARTHS

To model ARTHS neurodevelopment *in vitro*, we derived iPSCs from ARTHS patients and matched controls. All 5 ARTHS patient-derived iPSC lines (2 early-truncating, 3 late-truncating) were from patients previously reported in the literature (**Fig. 2A**). Control lines were derived from unaffected sex-matched individuals. All lines were differentiated in parallel batches containing at least one line from each group, with 3 independent differentiations per line.

**Fig. 2.**
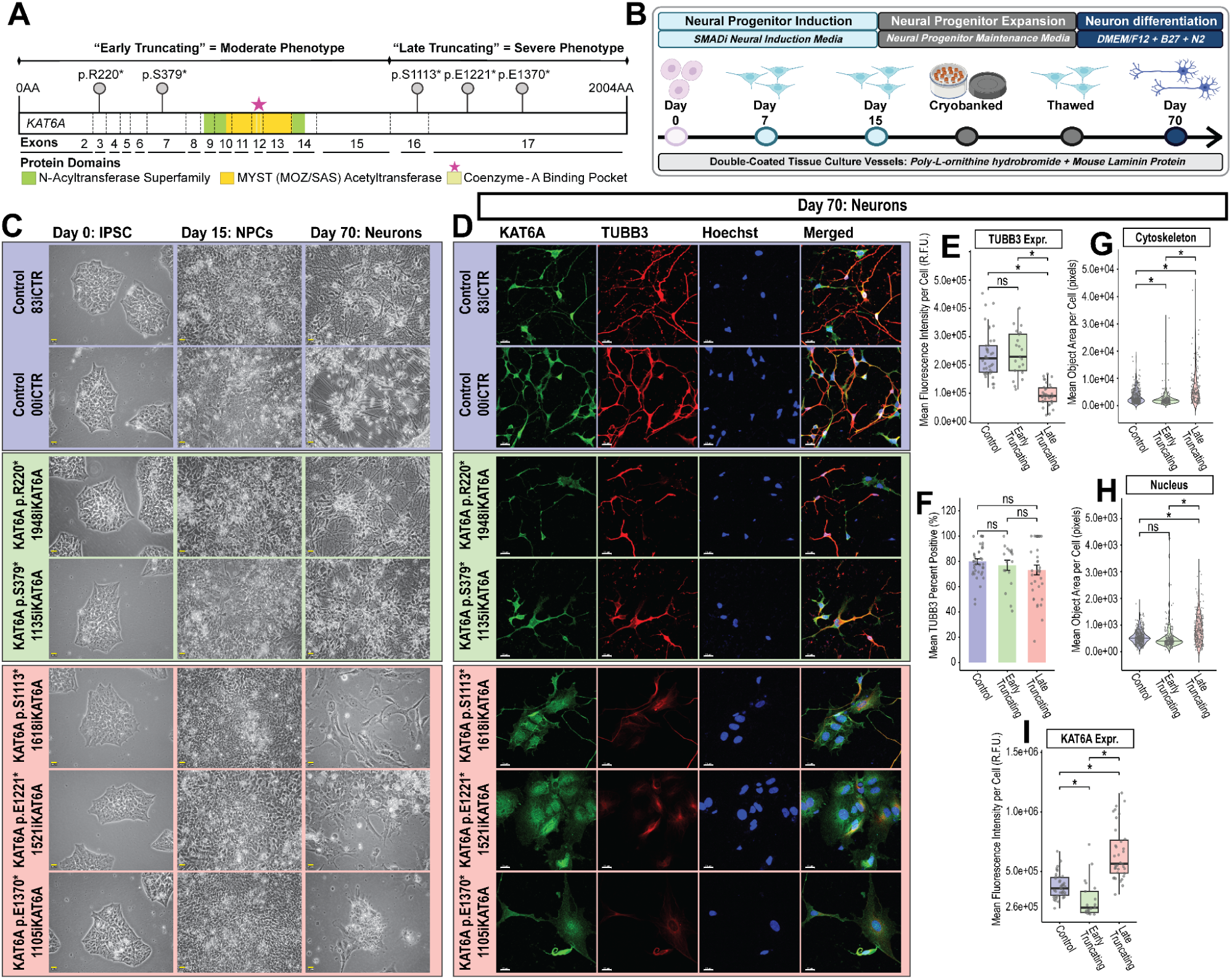
Early- and late-truncating KAT6A variants cause opposite changes in neuronal morphology and KAT6A protein abundance. **A**, Schematic of pathogenic *KAT6A* variants in ARTHS patient-derived iPSCs: 2 early-truncating variants (exons 1–15), 3 late-truncating variants (exons 16–17), and 4 matched controls. **B**, Timeline of 70-day iPSC-to-neuron differentiation protocol, demarcating key developmental stages (iPSC, NPC, neuron). **C**, Representative bright-field images of key timepoints show progressive morphological changes during differentiation. Late truncating cultures show visibly enlarged cells relative to control and early-truncating cultures at day 70. Scale bar, 20 µm. **D**, Immunofluorescence (IF) images of day 70 cultures stained for *KAT6A* (green), *TUBB3* (red, neuronal marker), and Hoechst (blue, nuclear). Scale bar, 20 µm. **E-I**, Quantitative IF results: statistical analysis of cell-count normalized values (mean per cell: fluorescent intensity, fluorescent area, or % TUBB3 positive) was performed using unpaired t-tests followed by Benjamini-Hochberg (BH) correction, comparing the 3 genotypes (control, early, late) where * = p-adj < 0.05 and ns = p-adj > 0.05. **E**, Quantification of mean TUBB3 expression per cell in day 70 cultures: late-truncating neurons express significantly less TUBB3 than early-truncating and control neurons (p-adj<0.05); individual points overlying plot = individual images from 9 biologically-independent cell lines with replicates: 4 control, 2 early, 3 late lines. **F**, Quantification of percent TUBB3-positive in day-70 cultures shows no significant differences across groups; individual points overlying plot = individual images from 9 biologically independent cell lines with replicates: 4 control, 2 early, 3 late lines. **G**, Quantification of mean cytoskeleton area (TUBB3) per cell in day-70 cultures: early-truncating neurons have significantly smaller cytoskeletons than controls; late-truncating cells have significantly larger cytoskeletons (p-adj<0.05); individual points overlying plot = individual cells from 9 biologically-independent cell lines with replicates: 4 control, 2 early, 3 late lines. **H**, Quantification of mean nuclei area (hoechst) per cell in day-70 cultures. Late-truncating day-70 cells have significantly larger nuclei than early-truncating and control day-70 cells (p-adj<0.05); individual points overlying plot = individual cells from 9 biologically-independent cell lines with replicates: 4 control, 2 early, 3 late lines. **I**, Quantification of mean full-length KAT6A protein expression per cell in day-70 cultures using C-terminal specific antibody: early-truncating variants cause *KAT6A* loss-of-function (69.5% of control, p-adj<0.05), while late-truncating variants cause gain-of-function (172% of control, p-adj<0.05); individual points overlying plot = individual images from 9 biologically-independent cell lines with replicates images per each line differentation: 4 control, 2 early, 3 late lines.

All iPSC lines showed equivalent pluripotency and proliferation prior to differentiation, and all acquired NPC identity by days 7 and 15, with both truncating classes proliferating more than controls (**Supplementary Notes 1–2, Extended Data Figs. 2A–L, 3A–H**). Using modified published protocols ^20–22^ we established a 70-day workflow from iPSCs through NPCs to neurons (**Fig. 2B**). No morphological differences were detected across groups until day 70 *in vitro* (hereafter day-70 cells), when late-truncating cells appeared enlarged and lacked projections relative to controls (**Fig. 2C**).

After 30 days in neuron differentiation media, late-truncating cultures expressed less TUBB3 than controls and early-truncating cells (p-adj = 1.15 × 10⁻¹³; **Fig. 2D–E**), although all groups contained a similar percentage of TUBB3-positive cells (Fig. 2F, all p-adj > 0.05), indicating that reduced TUBB3 does not reflect a failure of neuronal induction. Day-70 late-truncating cells also expressed almost no MAP2 (p-adj = 2.90 × 10⁻¹³) and retained Ki67 immunoreactivity, but expressed no PAX6, and so had not reverted to an NPC identity (**Extended Data Fig. 3I–K**). Early- and late-truncating *KAT6A* variants therefore both disrupt neuronal differentiation, with variant class determining its direction.

### Variant position regulates neuron morphology via differential KAT6A protein abundance

Early- and late-truncating neuronal cultures exhibited opposing morphological phenotypes. At day 70, early-truncating cells showed significantly smaller cytoskeletons despite normal TUBB3 expression, while late-truncating cells displayed significantly larger cytoskeletons and nuclei with reduced TUBB3 expression (**Fig. 2G-H**). To determine whether differential KAT6A protein abundance underlies these morphological differences, we performed quantitative IF on neurons using an antibody targeting the KAT6A C-terminal region (residues 1330–1404). This antibody specifically recognizes full-length *KAT6A*, since early-truncating variants lack the complete epitope and late-truncating variants (except p.E1370*) are truncated upstream of this region.

Early-truncating variants significantly reduced KAT6A protein to 69.5% of control levels, while late-truncating variants elevated it to 172% (**Fig. 2I**), indicating loss- and gain-of-function mechanisms, respectively. *KAT6A* localized to both the nucleus and the cytoplasm, consistent with known subcellular distribution^23^. These findings establish that variant-specific *KAT6A* protein dosage directly determines neuronal morphology, mechanistically linking truncation position to cellular phenotype through differential protein abundance.

### Late-truncating KAT6A variants are shifted to immature neuronal transcriptional state

To assess the molecular basis of the abnormal morphology in late-truncating day 70 cells relative to the other groups, we performed RNA sequencing (RNA-seq) across all groups at day 0, day 7, day 15, and day 70 *in vitro* (timepoints in **Fig. 2B**). Principal component (PC) analysis revealed the RNA-seq data separated along PC1 with iPSC (day 0) and neuronal (day 70) samples at opposing poles of PC1. (**Fig. 3A**). The day 70 cultures harboring late-truncating variants clustered away from control and early-truncating day-70 neuronal samples and towards NPC samples indicating transcriptional immaturity.

**Fig. 3.**
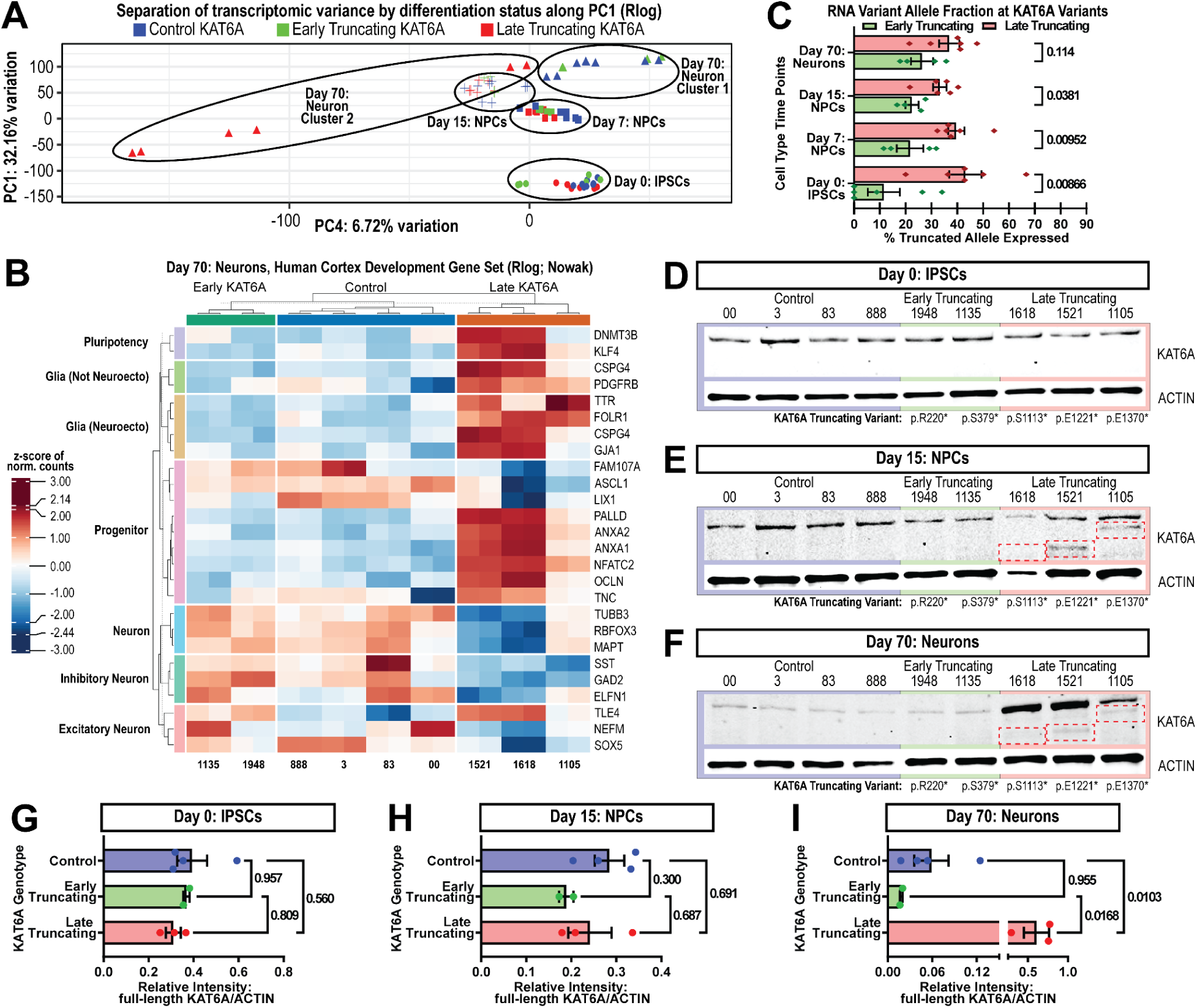
Variant position drives cell type-specific *KAT6A* protein abundance through differential nonsense-mediated decay during neuronal differentiation. **A**, Principal component (PC) analysis of regularized log (rlog)-transformed RNA-seq gene counts across a 70-day differentiation model identified stratification of samples by differentiation stage along PC1: “Day 0: iPSC” = ⬤, “Day 7: NPCs” = ▪, “Day 15: NPCs” = ✚, and “Day 70: Neuron” = ▲. Individual points = individual libraries from 9 biologically-independent cell lines with replicates: 4 control, 2 early, 3 late lines. **B**, Unsupervised hierarchical clustering of gene-wise z-scored, rlog-transformed RNA-seq counts from day-70 cells. Only genes (rows) related to cortical development and pluripotency are plotted. Individual columns = individual libraries from 9 biologically-independent cell lines with replicates: 4 control, 2 early, 3 late lines. **C**, RNA variant allele fraction analysis of truncating *KAT6A* variants across in vitro differentiation (n = 2 early- & 3 late-lines with replicates per timepoint). Early-truncating samples show significantly lower expression of the truncating KAT6A allele compared to late-truncating samples, consistent with NMD rules; bar plots show mean ± SEM % truncated allele expressed in early- vs late-truncating, compared within each timepoint using two-sided Mann-Whitney U test: “Day 0: iPSC” = 11.56 ± 6.16% vs 43.13 ± 6.28% with p = 0.008658; “Day 7: NPC” = 21.70 ± 5.15% vs 39.56 ± 3.15% with p = 0.009524; “Day 15: NPC” = 22.49 ± 2.59% vs 33.21 ± 2.65% with p = 0.038095; “Day 70: neuron” = 26.35 ± 4.25% vs 36.87 ± 3.89% with p = 0.114286; across all 4 timepoints = 19.53 ± 2.79% vs 38.19 ± 2.11%. Individual points = individual libraries. **D-F**, Western blotting assessment of full-length KAT6A protein expression (∼225 kDa) in whole cell lysate from “Day 0: iPSCs”, “Day 15: NPCs”, “Day 70: Neurons”; red dashed boxes indicate truncated KAT6A protein from late-truncating patient with p.S1113* (∼122 kDa), p.E1221* (∼134 kDa), p.E1370* (∼151 kDa). Lanes = 9 biologically-independent cell lines: 4 control, 2 early, 3 late lines. Loading control = actin (∼42 kDa). **G-I**, Quantification of full-length KAT6A band intensity, normalized to actin, from the western blots shown in D-F; truncated versions of KAT6A (red dashed boxes in D-F) were excluded from quantification. Bars show mean relative full-length KAT6A intensity ± SEM, where individual points represent quantification of individual lanes. In each blot quantification, the groups (control, early, late) were compared using a one-way ANOVA followed by Tukey’s multiple comparisons test (n = 4 control, 2 early, 3 late lines). Full-length KAT6A expression did not significantly differ between the groups at “day 0: iPSC” (ANOVA: F(2,6) = 0.59, p = 0.58) or “day 15: NPC” (ANOVA: F(2,6) = 1.38, p = 0.32). At “day 70”, late-truncating cells expressed significantly more full-length KAT6A compared to control (Tukey p = 0.010) and early (Tukey p = 0.017) cells, which did not differ from each other (p = 0.95); ANOVA: F(2,6) = 12.15, p = 0.0078.

Next, we examined RNA expression of genetic markers of cell types involved in human cortical development^24^ (**Fig. 3B**). Late-truncating neuronal cultures retained high RNA expression of genes associated with pluripotency and neuroprogenitor cells and low RNA expression of neuronal genes relative to control and early-truncating neuronal cultures. We stained neuronal cultures for SOX10 and GFAP to determine whether late-truncating cultures acquired an alternative lineage identity during neuronal differentiation but found no detectable SOX10 or GFAP expression across any group (**Extended Data Fig. 3L-M**). Late-truncating day-70 cells therefore retain an immature transcriptional profile without adopting NPC, glial, or neural crest identity (**Fig. 2D-I, 3A-B; Extended Data Fig. 3I-M**).

### NMD drives allele-specific expression of KAT6A in early- and late-truncating samples

Because NMD efficiency decreases with differentiation^7^, we examined *KAT6A* transcript allele fractions across cell differentiation. We analyzed RNA-seq data from control and patient-derived cell-lines harboring truncating *KAT6A* variants at day 0, 7, 15, and 70 of *in vitro* neuron differentiation (**Fig. 3C**). Transcripts carrying early-truncating variants were significantly depleted relative to late-truncating variants. Across all timepoints, the mean variant allele fraction of early-truncating and late-truncating samples was 19.53 ± 2.79% and 38.19 ± 2.11%, respectively; consistent with active NMD-mediated degradation of early-truncating transcripts.

Western blotting with an N-terminal antibody (residues 81–179; **Fig. 3D–F**) detected truncated KAT6A in all three late-truncating lines at days 15 and 70 but not at the iPSC stage, and in no early-truncating sample. Full-length *KAT6A* abundance did not differ across groups in iPSCs or NPCs (**Fig. 3G–H**). At day 70, full-length *KAT6A* protein differed across the three groups (p = 0.0078; **Fig. 3I**), with late-truncating cells expressing more than controls (Tukey p = 0.010) and early-truncating cells (Tukey p = 0.017); control and early-truncating cells did not differ from each other (Tukey p = 0.95). Whereas per-cell immunofluorescence showed a reduction (**Fig. 2I**). The two measures use different denominators: protein per cell versus per µg lysate, and early-truncating neurons have reduced cell size and cytoskeletal content **(Fig. 2D–H)**, such that reduced KAT6A per cell is compatible with unchanged KAT6A per unit protein.

Early-truncating variants undergo NMD and reduce KAT6A dosage; late-truncating variants escape NMD, sustain truncated protein, and increase full-length KAT6A. KAT6A abundance therefore departs from control in opposite directions in the two classes — the first of several measures to do so.

### Variant-specific differential KAT6A protein dosage perturbs gene regulation of cell fate modulators and their targets during in vitro neurodevelopment

Multi-omic profiling confirmed variant-class-specific *KAT6A* protein differences (**Fig. 4A**; consistent with **Fig. 3**) and revealed widespread perturbation of chromatin accessibility and gene expression across neurodevelopmental loci in both variant classes (**Fig. 4A–T**, **Extended Data Figs. 4–5**).

**Fig. 4.**
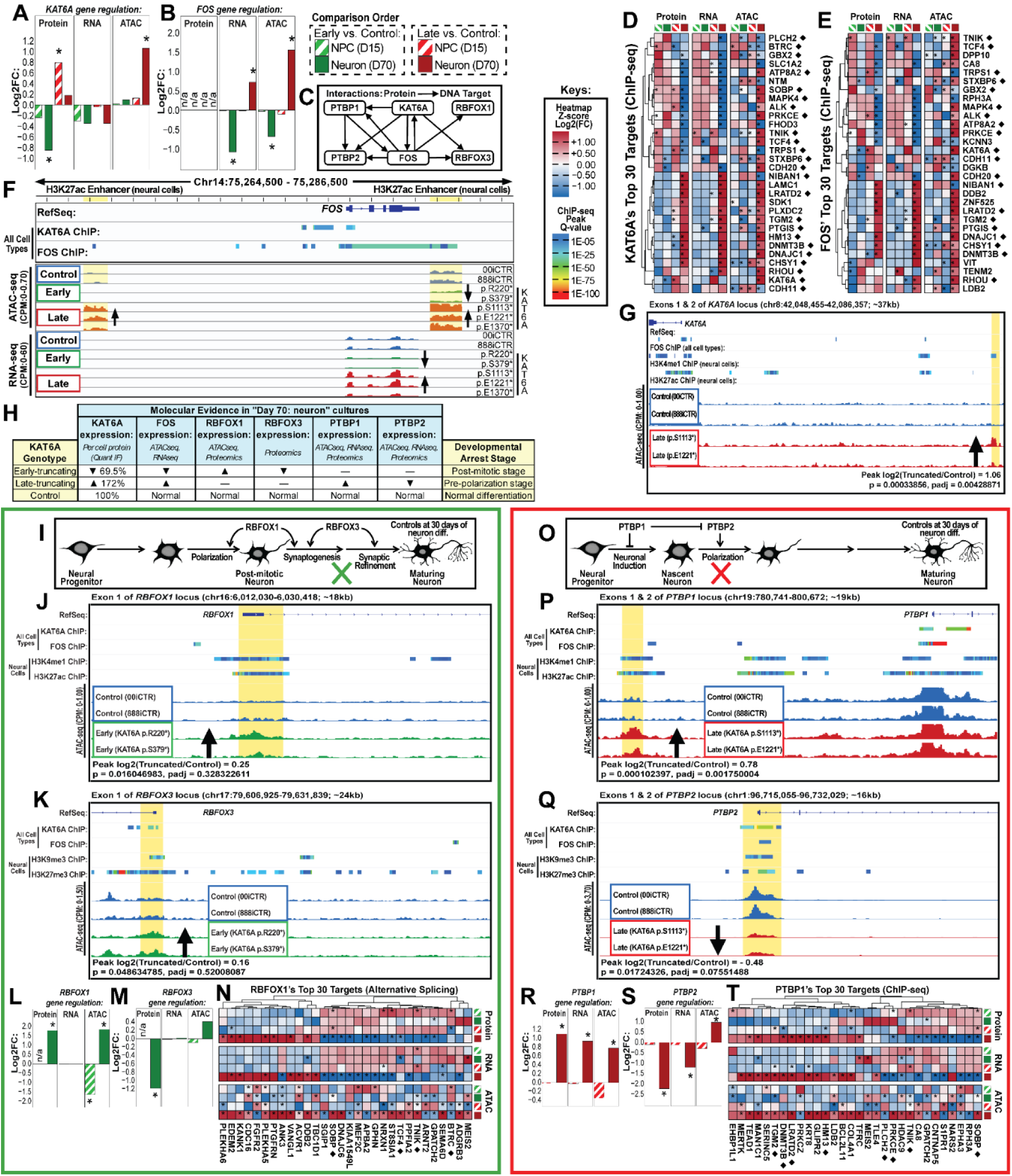
Opposite *KAT6A* dosage drives divergent neuronal differentiation via rewiring of the *KAT6A-FOS-RBFOX-PTBP* axis. **A-B**, Gene regulation plots of KAT6A and FOS. **C**, Gene regulatory network linking *KAT6A* dosage to divergent neuronal differentiation in early- and late-truncating variants. Nodes represent genes with significant differential chromatin accessibility (ATAC-seq) or expression (RNA-seq, proteomics); edges represent ChIP-seq interactions. **D-E**, Heatmaps of multi-omic dysregulation across the top 30 *KAT6A* and *FOS* ChIP target genes, respectively. **F**, Genomic tracks at the *FOS* locus in early- and late-truncating day-70 neurons relative to controls. Upper tracks: *KAT6A* and *FOS* ChIP-seq peaks (all cell types, ChIP-Atlas) and activating histone mark (H3K27ac) ChIP-seq peaks from neural cells. Lower tracks: differential ATAC-seq accessibility and RNA-seq expression. Yellow shaded regions mark *FOS* enhancer-overlapping ATAC-seq peaks with significantly reduced accessibility in early-truncating and increased accessibility in late-truncating cells. ATAC-seq signal outside significant peaks is not shown. **G**, Genomic tracks at the *KAT6A* 5’ region in late-truncating day-70 neurons relative to controls. Upper tracks: *FOS* ChIP-seq peaks (all cell types) and activating histone marks (H3K4me1, H3K27ac; neural cells). Yellow shaded region marks a significantly open ATAC-seq peak in late-truncating cells overlapping H3K4me1; DESeq2 statistics shown beneath ATAC-seq tracks. **H**, Summary table of significant multi-omic dysregulation in the *KAT6A*-driven gene regulatory network (**Panel C**) in early- and late-truncating day-70 neurons relative to controls. **I**, Schematic of the early-truncating variant gene regulatory network. At day 70, early-truncating neurons show elevated *RBFOX1* and reduced *RBFOX3* protein, consistent with developmental arrest at a post-mitotic stage (green X). Original artwork by A.A.N. **J-K**, Genomic tracks at the 5’ regions of *RBFOX1* (**J**) and *RBFOX3* (**K**) in early-truncating day-70 neurons relative to controls. Upper tracks: *KAT6A* and *FOS* ChIP-seq peaks (all cell types) and activating (H3K4me1, H3K27ac) or repressing (H3K9me3, H3K27me3) histone mark ChIP-seq peaks from neural cells. Yellow shaded region in **J**: open ATAC-seq peak in early-truncating cells overlapping activating histone marks. Yellow shaded region in **K**: open ATAC-seq peak in early-truncating cells overlapping *KAT6A* and repressing histone marks. **L-M**, Gene regulation plots of RBFOX1 and RBFOX3 in early-truncating day 15 and day 70 **N**, Heatmap of multi-omic dysregulation across the top 30 *RBFOX1* ChIP target genes. **O**, Schematic of the late-truncating variant gene regulatory network. At day 70, late-truncating neurons show elevated *PTBP1* and reduced *PTBP2*, consistent with developmental arrest at a nascent pre-polarization stage (red X). Original artwork by A.A.N. **P-Q**, Genomic tracks at the 5’ regions of *PTBP1* (**P**) and *PTBP2* (**Q**) in late-truncating day-70 neurons relative to controls. Upper tracks: *KAT6A* and *FOS* ChIP-seq peaks (all cell types) and activating (H3K4me1, H3K27ac) or repressing (H3K9me3, H3K27me3) histone mark ChIP-seq peaks from neural cells. Yellow shaded region in **P**: significantly open ATAC-seq peak in late-truncating cells overlapping activating histone marks. Yellow shaded region in **Q**: significantly closed ATAC-seq peak in late-truncating cells overlapping *KAT6A*, *FOS*, and repressing histone marks. **R-S**, Gene regulation plots of PTBP1 and PTBP2. **T**, Heatmap of multiomic dysregulation across the top 30 target genes of PTBP1. **A-B, D-E, L-N, and R-T**, All gene regulation plots displaying results from proteomic mass spectrometry (Protein), RNA-seq (RNA), and ATAC-seq (ATAC) — log₂(FC) is calculated as log₂(truncating/control), with significant hits marked with an asterisk; for RNA-seq and ATAC-seq significant genes or peaks are those with a p-adjusted < 0.05 and an absolute value of log₂(FC) > 0.58; for proteomic mass spectrometry significant genes are p-value < 0.05. Four comparisons shown: early-truncating vs. control (green) and late-truncating vs. control (red) across day 15 NPCs (striped) and day 70 cultures (solid). **D-E, N, and T**, Heatmaps display z-score of log₂(fold change) that is calculated per gene across 4 comparisons for each individual datatype (Protein, RNA, ATAC). Genes names with ◆ are amongst KAT6A’s top 30 ChIP-seq targets. **F, J-K, and P-Q**, ChIP-seq tracks show all available peak information from ChIP-Atlas, displaying only significant peaks with a Q-value < 0.00001 (MACS2).

*FOS*, encoding c-Fos, a leucine zipper transcription factor subunit of the AP-1 complex, emerged as the sole convergent node across chromatin and transcriptome levels, identified by systematic filtering for genes with concordant bidirectional dysregulation across connected-omic modalities and cell types (see Methods), significantly downregulated in early-truncating neurons and upregulated in late-truncating neurons relative to controls (**Fig. 4B, E, F**). Although c-Fos fell below mass spectrometry detection limits (**Fig. 4B**), consistent with its rapid induction kinetics and short half-life^25^, motif enrichment analysis across differential ATAC-seq peaks and differentially expressed genes revealed AP-1 complex binding sites (c-FOS, JUN, ATF, and MAF families) enriched at oppositely dysregulated loci in both variant classes, supporting a functional difference in FOS transcriptional activity. KAT6A and FOS ChIP-seq data^26–30^ revealed mutual binding at each other’s loci (**Fig. 4F–G**), corroborated by variant-class-specific chromatin accessibility changes at *FOS* enhancer sites^31^ in our ATAC-seq data (**Fig. 4F**), consistent with direct epigenetic regulation of *FOS* by *KAT6A*. Cross-referencing KAT6A and FOS ChIP-seq targets ^26–30^ with our neuronal gene regulatory data identified the RBFOX1/3-PTBP1/2 axis as differentially and inversely regulated between variant classes: *RBFOX1/3* dysregulation was specific to early-truncating neurons while *PTBP1/2* dysregulation was specific to late-truncating cells at day-70 of differentiation (**Fig. 4H–T**).

Only in early-truncating neurons were *RBFOX1* protein abundance and chromatin accessibility significantly increased, while *RBFOX3* protein was significantly decreased relative to controls (**Fig. 4L–M**). Differential ATAC-seq peaks at the *RBFOX1* 5′ region overlapped activating histone marks (H3K4me1, H3K27ac), while those at *RBFOX3* overlapped *KAT6A* binding sites and repressive marks (H3K9me3, H3K27me3) (**Fig. 4J–K**) — consistent with the opposing protein-level changes and developmental arrest at a post-mitotic stage (**Supplementary Note 4**; **Fig. 4I–N**).

In late-truncating neurons, *PTBP1* was significantly increased across all three modalities while *PTBP2* protein and RNA were significantly decreased relative to controls (**Fig. 4R–S**); neither gene was differentially regulated in NPCs. A peak of increased accessibility at the *PTBP1* 5′ region overlapped activating histone marks (H3K4me1, H3K27ac), while a peak of reduced accessibility at *PTBP2* overlapped *KAT6A*, *FOS*, and repressive histone marks (H3K9me3, H3K27me3)(**Fig. 4P–Q**), consistent with the opposing protein-level changes and with arrest at a nascent pre-polarization stage^32^ **(Fig. 3)**.

Twenty-four of the top 30 KAT6A ChIP targets appeared across the FOS, RBFOX1, and PTBP1 heatmaps (z-scored log₂FC; **Fig. 4D–E, N, T**), supporting KAT6A as the central regulatory driver.

These findings support a model in which divergent *KAT6A* dosage drives opposing *FOS* levels, disrupting distinct RNA-binding protein networks in a variant-class-specific manner to stall neuronal differentiation at opposite developmental checkpoints (**Figs. 2-4**, **Extended Data Figs. 4-5**).

### Variant-Specific KAT6A Dosage Controls Mitochondrial Physiology and Metabolic State via GRP75

*FOS* modulates the metabolic state of neural lineages ^33–36^ and is associated with metabolic dysfunction in neurological disorders^37^. Integrating RNA-seq and proteomic data from truncating *KAT6A* neural lineages identified metabolism and brain-health genes differentially expressed relative to controls (**Extended Data Fig. 5**). *KAT6A* ChIP targets accounted for 23 of 30 differentially abundant proteins in NPCs (76.7%) and 27 of 39 in neurons (69.2%), roughly half to two-thirds of which have known metabolic or neuronal roles (**Extended Data Fig. 5A, D**).

Seahorse XF Analyzer bioenergetic profiling across iPSCs, NPCs, and day-70 neurons (**Fig. 5A–I**, **Extended Data Fig. 6A–I**) revealed stage-dependent metabolic consequences: both variant classes showed convergent bioenergetic changes in iPSCs and NPCs, consistent with the shared developmental transition from glycolysis to oxidative phosphorylation in post-mitotic neurons,^38^ while at the neuronal stage early- and late-truncating variants caused divergent profiles — early-truncating neurons showed significantly reduced OCR, ECAR, and ATP production while late-truncating neurons showed the inverse (**Fig. 5G–I**). *GRP75* (*HSPA9*/*Mortalin*)^39,40^ dose-dependently regulates mitochondrial morphology and oxidative-stress sensitivity in neural lineages^41–45^. Relative to controls, early-truncating neurons had significantly reduced GRP75 and late-truncating neurons had significantly elevated GRP75 (**Fig. 5J-K**). Early-truncating neurons showed significantly reduced mitochondrial content and a significantly higher shape factor, while late-truncating neurons contained significantly larger and more abundant mitochondria (**Fig. 5L-N**).

**Fig. 5.**
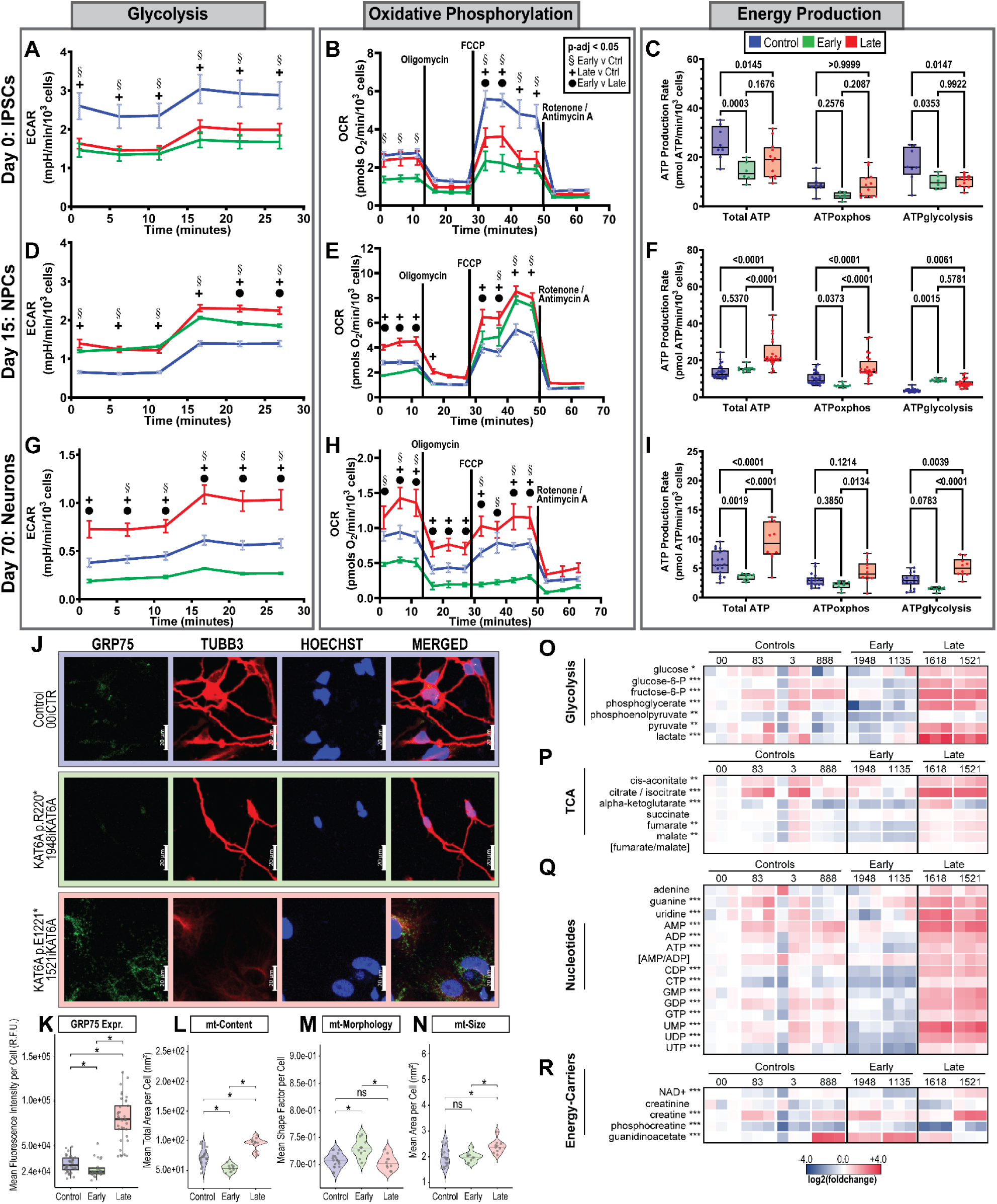
Early- and late-truncating KAT6A variants induce opposite metabolic phenotypes through inverse effects on mitochondrial dynamics. **A-I**, Seahorse XF analysis of mitochondrial bioenergetic profiles (oxygen consumption rate, OCR; extracellular acidification rate, ECAR) of **A-C** day 0 iPSC, **D-F** day 15 NPC, and **G-I** day 70 cultures from control (blue), early-truncating (green), and late-truncating (red) lines. At day 70, the two KAT6A truncating variant classes exhibit opposite bioenergetic profiles; early-truncating cells have significantly reduced OCR, ECAR, and ATP production reflective of a metabolic loss-of-function. Late-truncating cells have significantly elevated OCR, ECAR, and ATP production consistent with a metabolic gain-of-function (n= 9 lines). All Seahorse plots show mean ± SEM and significant differences among the three groups were assessed via two-way ANOVA followed by Tukey’s post-hoc multiple comparison test (p-adj < 0.05). Significance symbols for OCR and ECAR traces are: § = early vs. control; ✚ = late vs. control; ⬤ = early vs. late. **J**, Representative confocal images of day-70 neurons co-stained for GRP75 and TUBB3. Scale bar, 20 μm. **K-N**, Quantitative IF of neuron mitochondria: “mean per cell GRP75 expression” in panel K and “mean per cell mitochondrial physiology” in panels L-N; all comparisons are unpaired two-sided t-tests with Benjamini–Hochberg correction. **K**, Mean GRP75 fluorescence intensity per cell in day-70 cells, imaged via confocal (n = 4 control, 2 early-truncating, 3 late-truncating lines with replicates). Box plots show the mean expression per cell per image, with individual points = individual images. Relative to controls, GRP75 expression was significantly reduced in early-truncating cells (82% of control, p= 0.0363) and significantly increased in late-truncating cells (267% of control, p= 6.69E-15). **L-N**, Quantification of mitochondrial content (panel L, total mitochondrial area per cell), morphology (panel M, shape factor per cell), and size (panel N, mean mitochondrial area per cell) in day-70 neurons; n= 4 control, 2 early-truncating, 3 late-truncating lines with replicates. Violin plots show “mean per cell mitochondrial physiology”, with individual points = individual wells. Compared to controls, early-truncating cells had significantly reduced mitochondrial content (75% of control, p = 1.51E-4) and increased shape factor (103% of control, p = 3.35E-4). Compared to controls, late-truncating cells had significantly increased mitochondrial content (133% of control, p = 5.37E-6) and larger individual mitochondria (119% of control, p = 7.17E-3). **O-R**, Results of labeled glucose (U-¹³C₆) metabolite mass spectrometry: heatmaps of the log₂(FC) of total pooled metabolite levels in early-, late-truncating and control neuronal cultures; log₂(FC) is calculated as log₂ (individual sample value/average value of control group) where each metabolite value represents the sum of all its isotopomers after input normalization for cell number and trifluoromethanesulfonate. Heatmaps contain 29 of 75 differentially abundant metabolites: 7 glycolysis-related, 5 TCA-related, 13 nucleotide-related, 4 energy carrier-related; remaining 46 of 75 significantly differentially abundant metabolites are discussed in associated extended data. All significance tests were performed on normalized data with a one-way ANOVA; * p < 0.05, ** p < 0.01, *** p < 0.001.

U-¹³C₆ D-glucose tracing in day-70 neurons identified 75 differentially abundant metabolites across 40 pathways (**Fig. 5O–R; Extended Data Figs. 6J–N, 7A–G**). Late-truncating neurons showed significantly greater total metabolite abundance and higher labelled-glucose incorporation than early-truncating and control neurons; early-truncating neurons showed the inverse.

One exception was the glutamate, GABA, and glutamine cycle: late-truncating neurons utilized labeled glucose at significantly lower rates for the synthesis of glutamate, GABA, glutamine, α-ketoglutarate, and succinate relative to early-truncating and control neurons, yet had significantly greater total abundance of glutamate, GABA, and glutamine — with no difference in α-ketoglutarate or succinate (**Extended Data Fig. 7F–G**). This dissociation indicates that late-truncating neurons preferentially source glutamate from non-glucose carbon sources; in the absence of astrocytes in this *in vitro* system, the most likely exogenous source is glutamine supplied in the culture media^46^, consistent with the known capacity of neurons to utilize extracellular glutamine for glutamate synthesis.

The excess glutamate in late-truncating neurons may further amplify *FOS* expression through glutamate receptor signaling as positive feedback. We treated day-70 neurons with kainate, a glutamate receptor agonist^47^, and performed RNA-seq.

Kainate suppressed global protein-coding gene transcription in both variant classes relative to controls (**Extended Data Fig. 7H**), but selectively induced AP-1 target gene transcription in late-truncating neurons (**Extended Data Fig. 7I**), consistent with a larger functionally active glutamate reservoir and a hypersensitized glutamate receptor signaling response in these cells. Together with the KAT6A→*FOS* axis (**Figure 4**), these data suggest metabolic glutamate accumulation constitutes a secondary amplifying mechanism. Across mitochondrial dynamics, bioenergetics, and metabolite production, *KAT6A* variant-class dosage drives metabolic trajectories that diverge in direction rather than degree in neurons, establishing *KAT6A* as a dosage-sensitive regulator of neuronal metabolic state.

### Pharmacological repression of KAT6A HAT activity rescues pathogenic signatures in late-truncating neurons

To test whether pathogenic signatures in late-truncating neurons could be rescued by inhibiting KAT6A HAT activity, we treated late-truncating lines with PF-9363, a KAT6A/KAT6B competitive HAT inhibitor ^13,15^, or DMSO vehicle during neuronal differentiation (**Fig. 6A**). Treatment began on day 12 of the 30-day NPC to neuron differentiation protocol, coinciding with the timepoint at which all late-truncating cultures maintained proliferation in contrast to control cultures. After 18 days of treatment, we observed a morphological shift in the late-truncating cultures towards a more control-like appearance—smaller cell bodies with visible neurite extensions relative to DMSO-treated cultures(**Fig. 6B**)—and RNA-seq was performed to assess molecular rescue.

**Fig. 6.**
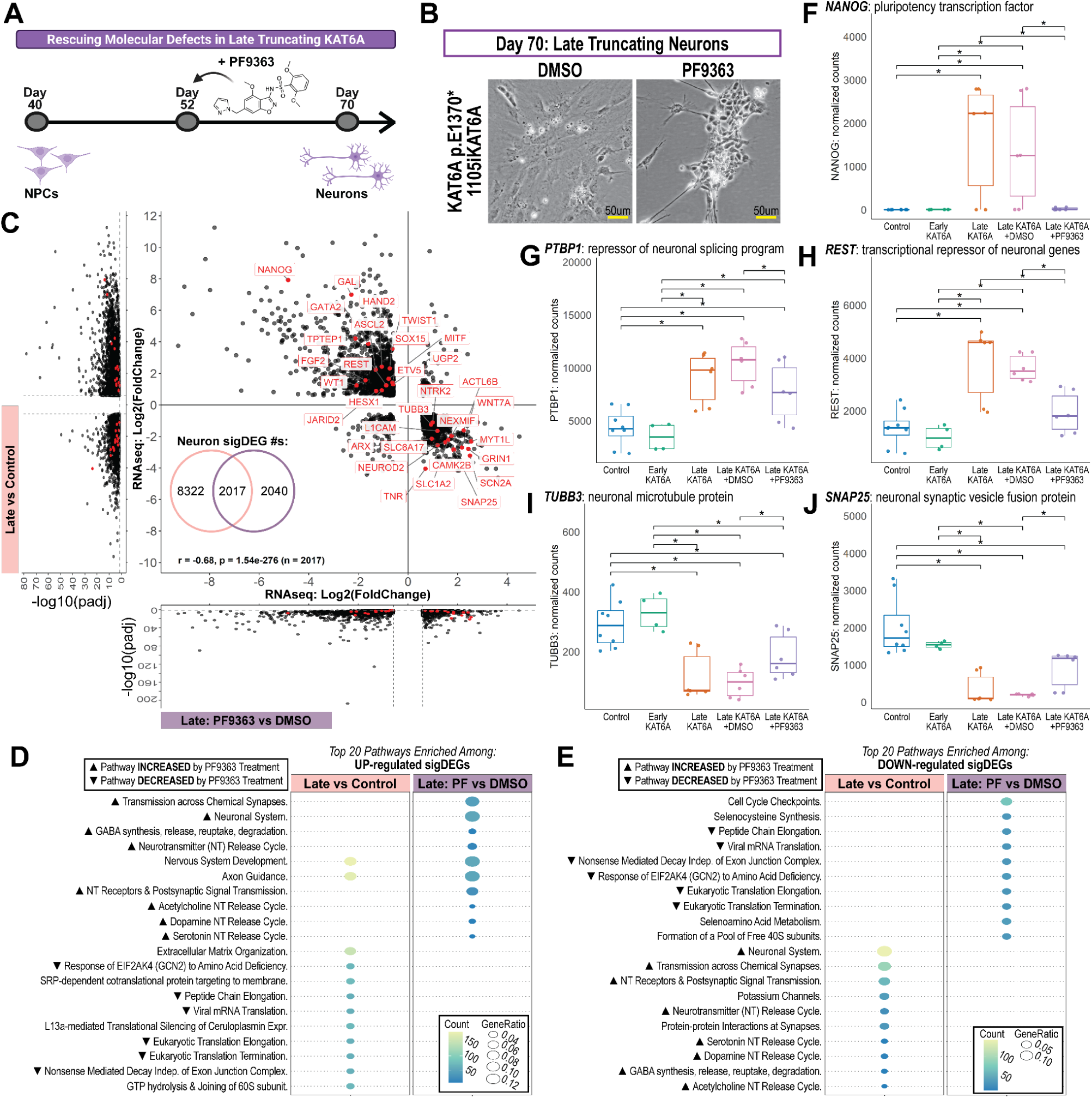
KAT6A/KAT6B competative HAT inhibitor, PF9363, rescues late-truncating pathogenic signatures and reveals therapeutic vulnerability. **A**, Overview of experimental design: late-truncating and control NPC lines were used to rescue molecular defects observed in day 70 cultures of late-truncating lines with PF9363 (1µM), a KAT6 HAT inhibitor from day 52 to day 70 *in vitro* during neuron differentiation. **B**, Representative bright-field images of late-truncating cultures at day 70 *in vitro* after 18 days of DMSO (left) or PF9363 (right) treatment. Scale bar, 50 µm. **C**, Significantly differentially expressed genes (sigDEGs) from “Late vs. Control” (left volcano) intersected with sigDEGs from “Late: PF9363 vs. DMSO” (bottom volcano); the 2,017 shared sigDEGs are plotted. Labeled genes have established roles in neurodevelopment. Genes in quadrants 1 and 3 are dysregulated in late-truncating day-70 neurons and reversed by PF9363 treatment. **D-E**, Top 20 pathways enriched among all upregulated or all downregulated sigDEGs from the two RNA-seq comparisons shown in panel C. Triangle symbols denote 14 enriched pathways rescued by PF9363 treatment: ▴ are neuro-related pathways downregulated in “Late vs. Control” and restored in “Late: PF9363 vs. DMSO”; ▾ are stress-related pathways upregulated in “Late vs. Control” & suppressed in “Late: PF9363 vs. DMSO”. **F-J**, Pairwise comparison of normalized gene counts from day 70 RNA-seq across neuronal fate repressor (*NANOG*, *PTBP1*, *REST*) and pro-neuronal (TUBB3, SNAP25) genes; asterisk denotes p < 0.05 in unpaired t-test with Benjamini–Hochberg correction. Two RNA-seq comparisons referenced throughout: late-truncating variants vs. unaffected controls (Late vs. Control) and late-truncating variants treated with PF9363 vs. DMSO (Late: PF9363 vs. DMSO).

To quantify rescue at the transcriptome level, we integrated results from two RNA-seq comparisons: “late-truncating vs. control” (**Fig. 4**, **Extended Data Fig. 4**) and “late+PF9363 vs. late+DMSO” (**Fig. 6A-B**). A log₂(FC) scatter plot of the 2,017 genes significantly differentially expressed in both comparisons revealed a negative correlation (Pearson r = −0.68, p = 1.54×10⁻²⁷⁶), reflecting directional reversal of gene expression between paradigms (**Fig. 6C**). Of these 2,017 genes, 90.1% showed log₂(FC) values in opposite directions (**Fig. 6C**). These results demonstrate that pharmacological KAT6 HAT inhibition reverses the transcriptional consequences of late-truncating *KAT6A* variants at the genome-wide level.

Pathway-level enrichment analysis of significantly up- and downregulated genes from both comparisons revealed a reciprocal pattern (**Fig. 6D-E**). In late-truncating neurons, stress-response pathways were enriched among upregulated genes and neurotransmitter-related pathways among downregulated genes relative to controls. PF-9363 treatment reversed this: stress-response pathways were enriched among downregulated genes and neurotransmitter-related pathways among upregulated genes in the “late+PF9363 vs. late+DMSO” comparison. This reciprocal enrichment pattern indicates pathway-level rescue of pathogenic molecular signatures in late-truncating neurons.

Consistent with this, PF-9363 treatment significantly decreased expression of neuronal fate repressor genes aberrantly elevated in late-truncating neurons (NANOG, PTBP1, REST; **Fig. 6F-H**) and significantly increased expression of pro-neuron genes aberrantly suppressed (TUBB3, SNAP25; **Fig. 6I-J**), reinforcing transcriptome-wide evidence of molecular rescue.

### KAT6A inhibition in control neurons recapitulates early-truncating signatures and identifies shared pathogenic pathways

Treating control NPCs with PF-9363 pharmacologically models *KAT6A* loss-of-function. Multi-omic integration identified 12 genes dysregulated across chromatin and transcriptome layers, most prominently *EGR1*, a FOS-interacting transcription factor^48^, whose enhancer showed reduced accessibility in both early-truncating and PF-9363-treated control neurons (**Extended Data Fig. 8A–C**). Twenty-four genes were differentially expressed in both loss-of-function paradigms, including *FOS* (**Extended Data Fig. 8D**). Comparative enrichment across all neuron RNA-seq comparisons identified 8 core pathways and 8 shared genes as candidate biomarkers, including *PTGIS*, previously dysregulated in ARTHS patient fibroblasts^49^ (**Extended Data Fig. 8E–H**). SPEAR multi-omic factor analysis^50^, excluding PF-9363 groups, identified a single factor separating late-truncating samples across all four omic layers and enriched for PF-9363-responsive genes (**Extended Data Fig. 9).** These results establish KAT6A HAT inhibition is a pharmacological strategy that reverses late-truncating pathogenic signatures at the genome-wide, pathway, and gene level, and demonstrate that comparative analysis using PF-9363-treated control and late-truncating neurons provides a systematic framework for identifying therapeutic targets and candidate biomarkers relevant to both truncating variant classes.

## DISCUSSION

Here we resolve the molecular mechanism by which truncation location drives divergent ARTHS clinical outcomes, establishing differential NMD-mediated *KAT6A* abundance as the critical determinant. The current clinical diagnostic and therapeutic target paradigm treats genetic diagnosis primarily on the gene level, with little consideration of how different variants drive divergent disease mechanisms. Our work challenges this reductionist view in a clinically actionable way. We demonstrate that mechanistic understanding of how variants disrupt protein function—not gene-level diagnosis alone—is essential for diagnostic and therapeutic strategy selection.

This principle is not new, with precedents in *SOX10*, *MPZ*, *MN1* ^8–12,51^— yet it has not reached clinical models or trial design. ARTHS shows the cost: patients with radically different severities are grouped as one entity, a distinction absent from OMIM. *KAT6B* shows the converse. Genitopatellar and Say-Barber-Biesecker-Young-Simpson syndromes were recognized as distinct before being collapsed into a single genetic spectrum ^52,53^ which may obscure whether variant position drives that distinction too, as it does in ARTHS — an open question with direct diagnostic consequences.

Our mechanistic dissection—showing early and late truncations cause opposite loss- and gain-of-function states via NMD—reveals this clinical heterogeneity as predictable from molecular mechanism, not unexplained genetic background. A severity continuum predicts that late-truncating variants amplify the changes seen in early-truncating cells. They do not. KAT6A abundance, oxygen consumption and extracellular acidification rates, ATP production, GRP75 levels, mitochondrial content, and *FOS* expression each move in opposite directions from control in the two variant classes rather than to different degrees in the same direction. Effect sizes are consistently larger in late-truncating cells, matching their greater clinical severity, but direction and magnitude are separable: it is inversion of sign across chromatin, transcript, protein, and metabolite layers that distinguishes two mechanisms from one graded defect. This has immediate diagnostic and therapeutic implications: variant position within the gene body predicts not just phenotype, but which therapeutic strategy will work. The NMD-based stratification we describe extends beyond KAT6A, across thousands of genes, though NMD efficiency varies by cell type and gene^54–56^. Yet this principle remains underutilized in rare disease diagnosis and drug development.

Our clinical validation using GestaltMatcher and DNA methylation episignature demonstrates that variant-level stratification produces clinically observable biomarkers—early and late-truncating patients have distinguishable facial phenotypes and blood methylation signatures. This suggests precision stratification is not merely mechanistically elegant but clinically actionable: GestaltMatcher and episignatures can identify which variant class a patient harbors, guiding therapeutic selection before molecular testing.

A second major finding reframes how chromatin regulators function in neurodevelopment. Rather than controlling a single downstream pathway, *KAT6A* dosage orchestrates a coordinated rewiring spanning gene regulation, neuronal cell fate, and cellular metabolism — apparent pleiotropy that emerges here as mechanistically integrated through a single developmental hub. *FOS* is the sole gene bidirectionally dysregulated across both variant classes at the chromatin and transcriptome levels, with KAT6A ChIP targets accounting for 69.2–76.7% of differentially abundant proteins in neurons and NPCs respectively, and mutual regulation of each other’s chromatin accessibility further linking the two through transcription factor activity, splicing programs, and mitochondrial metabolism suggests that apparent pleiotropy in other chromatinopathies may similarly reflect mechanistic integration around a small number of regulatory hubs.

KAT6A dosage-dependent control of mitochondrial physiology is accompanied by inverse GRP75-mediated dynamics. Early literature established that metabolic state changes during neurodevelopment, where progenitors shift from glycolysis to oxidative phosphorylation during differentiation ^57,58^. Our finding that variant-specific KAT6A dosage determines opposite metabolic trajectories— associated with *KAT6A* dosage-dependent differences in GRP75 abundance and downstream metabolic gene expression—suggests developmental stage-specific metabolic demands are actively programmed by chromatin regulators.

PF-9363, a competitive KAT6A/B HAT inhibitor developed as an oncology agent, rescues late-truncating pathogenic signatures. This raises a strategic question: can epigenetic drugs developed for cancer be repurposed for epigene-driven developmental disease? The answer is conditional on mechanism. Late-truncating ARTHS involves *excess* KAT6A activity (gain-of-function)—precisely what HAT inhibitors target. Early-truncating ARTHS requires *restored* KAT6A function—the opposite target. Our work establishes a principle: mechanistic understanding dictates whether oncology-derived epigenetic drugs are therapeutic or harmful. This has pragmatic implications for the field: clinical trials of epigenetic inhibitors in rare disease must stratify patients by mechanistic subtype, not gene-level diagnosis.

Several limitations and avenues for future work warrant attention. First, our cellular findings derive from five patient lines without isogenic controls, so line-intrinsic genetic background cannot be formally excluded. Two features of the study argue against it as the dominant explanation. Pharmacological KAT6A/B inhibition reverses the late-truncating phenotype within each line, establishing dependence on ongoing KAT6A activity rather than on fixed background. More importantly, the same variant-class boundary separates 63 patients by facial gestalt and distinguishes blood methylation profiles, demonstrating that the dichotomy exists at the ARTHS clinical phenotype level rather than as a property of the particular individuals from whom lines were derived. Isogenic knock-in models will nonetheless be required to establish causality for individual molecular phenotypes. Deep mutational scanning, validation in additional cell types or mouse models, direct measurement of splicing consequences, and functional manipulation of GRP75 will be required to extend these findings.

## AUTHOR CONTRIBUTIONS

A.A.N. and V.A.A. conceptualized the project and designed all approaches. A.A.N. and V.A.A. wrote and edited the manuscript with input from all authors. A.A.N. and Y.P. generated and analyzed the Biodock-related data -- in addition to culturing cells. T.C.H. and J.M.L. performed the GestaltMatcher analysis and contributed facial portraits from published ARTHS cases. A.A.N., V.A.A., & S.M. recruited new ARTHS cases to the GestaltMatcher cohort, contributing patient facial portraits to the analysis. A.B., S.C., and R.W. generated the DNA methylation data and performed the episignature analysis. A.A.N., V.A.A., I.V., E.T., A.P., D.L., L.P., J.M.P., & J.D.O.E. recruited new ARTHS cases to the episignature cohort, contributing patient DNA extracts to the analysis. A.A.N., A.K., and H.C. generated and analyzed the metabolite mass spectrometry data. J.F. and H.P. performed the multi-omic integration with SPEAR. A.A.N., N.M., S.A., and W.E.L. generated and analyzed the kainate-related data. A.A.N., V.P., and J.A.W. generated and analyzed the proteomic mass spectrometry data. A.A.N., L.S., and C.B. generated and analyzed the mitochondrial bioenergetics and morphology data from seahorse and high-content imaging, respectively. A.A.N. and M.S. generated the ATAC-seq data. L.T.U. and W.E.L. provided supervision and subject-matter expertise in neuroscience and stem cell biology. A.A.N. performed all remaining experiments and generated/analyzed all remaining data not specifically mentioned above.

## FUNDING

A.A.N. was supported by the National Institute of Neurological Disorders and Stroke (NINDS) under award number F31NS141668, the Eli and Edythe Broad Stem Cell Training Fellowship, and the Eugene V. Cota-Robles Fellowship. V.A.A. was supported by the Keck Foundation Junior Faculty Award, the KAT6 Foundation, and the Rose Hill Foundation. This work was also supported by a California Institute for Regenerative Medicine (CIRM) grant awarded to W.E.L. under the award number DISC0-14519.

## ACKNOWLEDGEMENTS

We would like to thank the families and the KAT6 foundation for their encouragement and support of this research. We would also like to acknowledge the following core facilities for providing their technical expertise in generating some of the data from this work: the UCLA Technology Center for Genomics and Bioinformatics (TCGB) prepared and sequenced the kainate-related RNA-seq libraries, the UCLA Mitochondria and Metabolism Core assisted in performing the seahorse and mitochondrial morphology analyses, and the UCLA Proteome Research Center (PRC) performed the proteomic mass spectrometry.

## DECLARATION OF INTEREST

The following declarations of interest are made by the authors: W.E.L. is a founder and shareholder of Pelage Pharmaceuticals, Sardona Therapeutics, and Cellio Biotechnology. H.R.C. is a founder of Pelage Pharmaceuticals. The work presented here was not supported by any of these companies. The remaining authors report no conflict of interest.

## DATA AVAILABILITY

Sequencing data is deposited in the Gene Expression Omnibus (GEO) and will be made public upon publication. Raw proteomics mass spectrometry data is deposited in the MassIVE repository and will be made publica upon publication. The raw individual-level clinical data reported in this study cannot be deposited in a public repository due to privacy, legal, and ethical restrictions/regulations. Requests for additional information, data, resources, and/or reagents should be directed to, and will be fulfilled by, the corresponding author.

## CODE AVAILABILITY

This study did not generate new custom code or original software algorithms. As detailed in the Methods, all computational/statistical analyses in this study were performed using standard workflows and publicly available software tools.

## SUPPLEMENTARY FILES

**Supplementary tables 1-65**, containing experimental/statistical results for all figures and all extended data figures, are provided in a single file and are cited in numerical order in the methods section. Titles and legends for **supplementary tables 1-65** are also provided as a single file; legends contain information like sample sizes, statistical tests applied, and lines used - among other pertinent details.

## SOURCE DATA

Unprocessed and uncropped western blot images are provided for the western blot results shown in **figure 3**.

## METHODS

### 1. Research participants

This study includes a total of 79 ARTHS patients recruited across three cohorts included in either the GMDB, episignature, or *in vitro* iPSC analyses. Patients were recruited and consented in accordance with their respective Institutional Review Board and/or Ethics Committee approval. All 79 ARTHS patients were affected by heterozygous dominant truncating KAT6A mutations -- encompassing 41 males (13 early, 28 late) and 38 females (13 early, 25 late). Their deidentified metadata is provided and all patient variants are in reference reported using GR38 human reference genome (**Supplementary Table 1**).

### 2. GestaltMatcher: facial feature analysis

We used GestaltMatcher ^1^ to analyze frontal facial photographs from 63 individuals with KAT6A truncating variants, comprising 17 early-truncating and 46 late-truncating cases. The dataset included 49 cases extracted from 17 previous published reports ^2–18^ and 14 unpublished patient cases contributed by KAT6A patient groups and collaborators to the GestaltMatcher Database (GMDB)^19^. This dataset is based on published photographs and additional unpublished cases where written informed consent for publication of identifiable facial photographs was obtained from all participants or their legal guardians. This study was conducted in accordance with the Declaration of Helsinki. Each image was encoded into a 12-dimensional facial descriptor vector (FDP) using an ensemble of deep models with test-time augmentation to enhance stability ^20^. Cosine distance between FDPs served as the dissimilarity metric; lower values indicate higher facial similarity within the clinical face phenotype space (CFPS). The overall workflow is shown in **Fig. 1B**. Three complementary analyses were performed: t-SNE visualization, cohort-level distance-based comparison with PPV estimation, and image-level leave-one-out linear SVM classification ^21^. First, two-dimensional t-SNE visualization was used to inspect the spatial distribution of early- and late-truncating cases in CFPS (**Fig. 1C**). Second, cohort-level inter-group analysis was used to quantify facial separation between the early- and late-truncating groups. For this analysis, we performed 100 random subgroup samplings from each group and calculated the mean pairwise cosine distance between the sampled early and late subcohorts for each iteration (**Fig. 1D**).

For benchmarking, we used pooled same- and different-syndrome reference distributions generated from a GMDB reference set of 1,499 images from 1,182 individuals across 321 syndromes, none of which were used for model training. These reference distributions included: (1) a same-syndrome control, in which both sampled cohorts originated from the same syndrome, and (2) a different-syndrome control, in which the sampled cohorts originated from different syndromes. ROC analysis using mean pairwise cosine distance as the discrimination measure yielded a predefined dissimilarity threshold of c = 0.8963 by maximizing the Youden index, corresponding to a sensitivity of 0.9035 and specificity of 0.8024. For inter-group comparisons, if more than 50% of sampled mean distances exceeded c, this was interpreted as evidence that the two cohorts showed facial separation relative to the reference distributions.

To further quantify the distinctiveness of the early- and late-truncating groups, we calculated PPV_range using the previously established framework. The observed distance range was defined by the minimum and maximum of the 100 sampled early–late subgroup mean distances. Sensitivity_in_range was defined as the probability that distances from the different-syndrome reference distribution fell within this observed range, whereas specificity_out_of_range was defined as the probability that distances from the same-syndrome reference distribution fell outside this range. PPV_range was then calculated as: “ *PPV = sensitivity × p / [sensitivity × p + (1 − specificity) × (1 − p)]”* where p denotes the pretest probability. As in the original framework, we assumed p = 0.5 to represent no prior information favoring either same- or different-syndrome origin.

Third, we performed image-level leave-one-out linear support vector machine (SVM) classification to evaluate whether early- and late-truncating cases could be distinguished in the facial embedding space (**Supplementary Table 2**). Because each image was represented by twelve descriptor outputs, we trained separate linear SVM classifiers for each descriptor index and averaged the resulting out-of-sample SVM decision scores across the twelve classifiers. Classification performance was assessed using accuracy, balanced accuracy, and area under the receiver operating characteristic curve (AUC). Positive and negative decision scores indicated classification relative to the learned SVM decision boundary (**Extended Data Fig. 1C**).

### 3. DNA methylation profiling and data processing

This research was approved by the Research Ethics Board at The Hospital for Sick Children (REB#1000038847) and UCLA IRB#11-001087. Genomic DNA for all samples were extracted from peripheral blood and bisulfite converted using the EpiTect Bisulfite Kit (EpiTect PLUS Bisulfite Kit, QIGEN). Sodium bisulfite-converted DNA was hybridized to the Illumina Infinium Human Methylation EPIC v 2.0 Beadchip, which interrogated > 930,000 CpG sites found across the human genome. Sample processing was performed at The Center for Applied Genomics (TCAG), The Hospital for Sick Children, Toronto, Canada.

Samples were analyzed using the SeSAMe Bioconductor package v. 1.22.2 ^22^ to preprocess the data, perform quality control, normalize Illumina probes using the Noob method, perform background correction and extract β-values. Standard quality control metrics were performed in SeSAMe, including median intensity QC plots and density plots. Problematic probes were removed including those with p-values greater than 0.05 in 25% of samples (n=40,438), mapping inaccuracies (n=190), probes near SNPs with minor allele frequencies above 5% (n=104,615), probes specific to either the X or Y chromosome and mitochondrial DNA (n=24,964), and probes found to exceed the signal-detection p-value threshold (n=50,209). In total, n=180,073 probes were filtered out of our EPIC v 2.0 analysis, leaving a total of n=757,617 probes remaining for the analysis.

### 4. DNA methylation signature derivation

We assessed differentially methylated sites in whole blood-derived DNA from n=4 individuals with truncating variants in the last two exons (exons 16 and 17) of *KAT6A*, as compared to a selection of n=8 sex- and age-matched typically developing controls. This signature discovery cohort of 4 late patients included 2 males and 2 females with a mean age of 7.70 ± 3.38 years old (range 3.8 – 12 years old). The 8 matched controls used for the discovery signature included 4 males and 4 females with mean ages of 7.66 ± 3.10 years old (range 3.6 – 12.2 years old). Early-truncating patients, used to assess specificity of late-truncating signature, included 4 males and 3 females with a mean age of 9.96 ± 12.71 years old (range 1 – 37.5 years old). For all samples, we applied the blood cell-type estimation tool in SeSAMe. Differentially methylated CpG sites were identified using the *Limma* regression modelling package using sex and blood-cell proportions as covariates. The thresholds for differentially methylated CpG sites were Benjamini-Hochberg adjusted p-value < 0.05 and a |Δβ| > 0.1. The Δβ value represents the difference in average DNAm (β) between groups. Principal component analysis (PCA) and hierarchical clustering graphs were generated using Qlucore Omics Explorer v.3.9 (QOE, www.qlucore.com). Internal validation of the model was performed using a leave-one-out cross-validation analysis (**Extended Data Fig. 1D**). For each of the samples analyzed as part of the DNAm signature creation (KAT6A late) or classified using that signature (KAT6A early), we provide scores as they appear on the SVM machine learning classification model (**Supplementary Table 3**). An additional selection of n=31 controls and n=7 samples from individuals with early-truncating KAT6A variants were also visualized on the PCA and heatmap to both validate signature sensitivity, and test if “early” truncating *KAT6A* variants clustered distinctly from “late” truncating variants.

### 5. Institutional approvals and human iPSC generation

This study was approved by the Institutional Review Board and the Embryonic Stem Cell Research Oversight (ESCRO) committees at UCLA. All biological samples were collected after informed consent (IRB#11-001087). Somatic cells, whole blood or dermal fibroblasts, were isolated from five biologically distinct ARTHS patients and submitted to the Cedars-Sinai Regenerative Medicine Institute’s Induced Pluripotent Stem Cell Core Production Facility (https://csbiomfg.com/ipsc-core/) for conversion into iPSCs using episomal reprogramming with the following factors: OCT3/4, SOX2, KLF4, L-MYC, SHP53, and LIN28.As a part of their reprogramming quality control, Cedars-Sinai performed G-band karyotyping, pluripotency immunocytochemistry, alkaline phosphatase staining, and IDEXX cell line authentication. Four biologically distinct age- and sex-matched control iPSC lines were purchased from this same core facility, and were generated using the same methodologies. Information for all iPSC lines is provided (**Supplementary Table 4**).

### 6. Maintaining and cryobanking iPSC cultures

All iPSC lines used in this study were grown under chemically defined, feeder-free conditions in 37℃ incubators supplemented with 5% CO_2_ at atmospheric O_2_ concentrations. All iPSC lines were maintained on Nunc or Corning 6-well culture plates (ThermoScientific #140675; Corning #3516) coated with recombinant human vitronectin protein at a final concentration of 0.50 ug/cm^2^ (Gibco #A14700) in complete Essential 8 Medium (complete E8; Gibco #A1517001). A complete media change was performed daily to prevent spontaneous differentiation. Plates were coated as follows: first stock vitronectin (0.5 mg/mL) was diluted 1/100 in DPBS without calcium or magnesium (Gibco #14190144), then diluted vitronectin was immediately dispensed into plates, followed by either overnight or 2-hour incubation at 37℃ before use. Stock vitronectin (0.5 mg/mL) was aliquoted into 1.5 mL polypropylene tubes and stored at −80℃.

All iPSC lines were passaged as clumps or single cells upon reaching 80-85% confluency using Versene (0.02% EDTA solution, Lonza #17-711E) or StemPro Accutase (Gibco #A1110501). To passage, cells were washed once with 1X DPBS (Fisher Scientific 14-190-144), before incubating with a lifting agent for ∼5 minutes at 37C. After incubating with lifting agent, cells were collected in sterile plasticware and lifting agent was quenched with media before pelleting and resuspending in complete E8 medium supplemented with Y-27632 dihydrochloride at a final concentration of 10 μM (Tocris Bioscience #1254); 10 μM Y-27632 was included in the media for the first 12-18 hours after passaging to ensure optimal survival. To create 1000x single-use stock, Y-27632 powder was reconstituted to 10 mM in sterile nuclease-free water (Invitrogen #10977023) then filter sterilized (PES membrane, 0.22 um; Fisher Scientific SCGP00525) before aliquoting on ice into 1.5 mL polypropylene tubes and storing at −80C. All cultures were routinely tested for mycoplasma contamination per manufacturer instructions (Lonza #LT07-318).

All iPSC lines were expanded and cryobanked as follows to create stable stock per line. Upon reaching ∼75-85% confluency, prior to harvest cells were pretreated with 10 μM Y-27632 for once hour to minimize death. Pretreated cells were then processed similar to passaging to obtain single cell suspensions in complete E8 media containing 10 μM Y-27632. To make a stable stock per line, single cell suspensions from multiple wells of the same line were pooled into a 50mL conical and counted twice times with an Invitrogen Countess II (Thermoscientific C10228) to obtain an average cell count. Single cell suspensions were aliquoted into cryovials (Thermoscientific #375418PK) at ∼2 million cells per 1 mL of 1X cryopreservation media consisting of complete E8 media containing 10 μM Y-27632 and 10% DMSO. Cryovials were stored at −80C in Mr. Frosty (Nalgene Cat no. 5100-0001) for ∼24 hours before transferring into a liquid nitrogen tank for long-term storage.

### 7. Inducing, maintaining, and cryobanking NPC cultures

Monolayer neural progenitor cells (NPCs) were generated from monolayer iPSCs using Stem Cell Technologies’ NPC workflow which entailed 15 days of neural induction followed by 10 days of expansion before cryopreservation; the NPC workflow we used in this publication is a modified version of a published STAR protocol ^23^, with the main difference being we induced for 15 days instead of 11 days to ensure patient-derived lines also were differentiated into NPCs.

To reduce batch effect and maximize detection of variant-specific differences, it is essential that all cell types and their predecessors be seeded at the same density at the same time or closely staggered by 1-2 days if >9 lines. First, iPSC vials were thawed ∼14-21 days prior to the start of NPC induction and were passaged at least twice upon reaching ∼80-85% confluency before use in the NPC induction protocol, and iPSC cultures had daily media changes with complete E8 media. For the first passage after thawing, iPSCs were re-seeded at a density of 200,000 cells per well on vitronectin-coated 6-well plates (Corning 3516) in E8 media containing 10uM Y-27632 (Tocris Bioscience #1254). Similarly, these iPSC cultures were passaged a second time after thawing and re-seeded at 150,000 cells per well of a 6-well plate. These iPSC plates were then used to seed for NPC induction upon reaching ∼80-85% confluency.

Prior to starting NPC induction or NPC expansion or neuron differentiation, double-coated tissue-culture-treated plates, flasks, or slides need to be prepared as follows. The first coating is always Poly-L-ornithine hydrobromide (PLO, Sigma P365550MG) and the second coating is always mouse laminin (Gibco 23-017-015). Undiluted PLO stock (5mg/mL) is prepared in water (Invitrogen 10977015) by rehydrating 50mg of PLO powder in 10mL of water, vortexing for 10 seconds, and then filter sterilizing the solution with a PVDF 0.22uM filter (MilliporeSigma SE1M179M6). The PLO stock solution is then aliquoted into sterile 1.5mL polypropylene tubes (Fisher Scientific 21-402-903) and store at −20C with a maximum of three thaws before discarding. While undiluted laminin stock (1mg/mL) is prepared by thawing and aliquoting on ice into single-use sterile 1.5mL polypropylene tubes then storing at −80C. These two undiluted stocks, and their diluted working solutions, are always handled on ice to prevent premature polymerization. Briefly, the PLO stock is diluted to 0.05 mg/mL in ice-cold water and dispensed onto plates at a final concentration of 5.0 ug/cm2 then incubated for a minimum of 2 hours or overnight at 37C. After PLO incubation, the PLO solution is aspirated and the plates are washed once with ice-cold water before adding diluted laminin. The laminin stock is diluted to 0.01 mg/mL in ice-cold water and added to plates at a final concentration of 1.0 ug/cm2 then incubated overnight at 37C.

On day 0 of NPC induction (0 DPI), iPSC plates were harvested using accutase, counted, and resuspended at 1,000,000 cells per mL in STEMDiff neural induction media containing dual SMAD inhibition (NIM; Stem Cell Technologies 08582) ^24^ then supplemented with 10uM Y-27632. This cell suspension was then plated on PLO+laminin-coated 6-well plates at 2,000,000 wells per well for ∼210,000 cells per cm2; after plating these cells were considered passage zero (P.0). A complete media change was performed with NIM, without Y-27632, throughout the 15-day NPC induction phase. On day seven post-NPC induction (7 DPI), a.k.a. one week after starting NPC induction, these cells were harvested using accutase, counted, and resuspended at 2,000,000 cells per mL in NIM supplemented with 10uM Y-27632. This cell suspension was then plated on PLO+laminin-coated 6-well plates at 1,900,000 cells per well for ∼200,000 cells per cm2; after plating these cells were considered passage one (P.1).

On day fifteen post-NPC induction (15 DPI), these cells robustly expressed the canonical NPC markers PAX6 and NESTIN, so NPC induction was terminated and NPC expansion was started at this timepoint to create a stable stock of NPC lines. This was done by harvesting NPCs using accutase, counting, and resuspending cells in STEMDiff Neural Progenitor Media (NPM; Stem Cell Technologies 05833) supplemented with 10uM Y-27632 – followed by plating on PLO+laminin-coated 6-well plates at 833,000 cells per well for ∼87,000 cells per cm2; after plating these cells were considered passage two (P.2). Similar as before, a complete media change was performed with NPM, without Y-27632, throughout the 10-day NPC expansion phase which is referred to as 25-days post-neural induction.

On day twenty-five post-NPC induction (25DPI), after 10 days in expansion media, NPCs were cryopreserved as follows. Cells were harvested using accutase, counted, and resuspended in NPM supplemented with 10uM Y-27632 such that upon mixing this cell suspension with 2X cryopreservation media, the desired number of cells is obtained in 1mL; e.g. if 20,000,000 cells are isolated for a line this pellet would be resuspended in 5mL of NPM+Y-27632 then mix with 5mL of 2X cryopreservation media for 2,000,000 cells per cryovial (Thermoscientific #375418PK) in 1mL of 1X cryopreservation media. The 2X cryopreservation media was created by preparing an 80% FBS (Avantor 97068-085) and 20% DMSO (Sigma D2438-50) solution -- followed by filter sterilization. To create a stable stock, NPC lines were banked at ∼5-8 million cells per cryovial in 1.0-1.2mL of 1X cryopreservation media, with at least 10-15 cryovials banked per line; these vials were labeled passage three (P.3). Similar to iPSC cryopreservation, cryovials were stored at −80C in Mr. Frosty (Nalgene Cat no. 5100-0001) for ∼24 hours before transferring into a liquid nitrogen tank for long-term storage.

### 8. Neuron differentiation

The NPC-to-neuron workflow used in this study is a modified version of a published spontaneous differentiation protocol ^25^, with the main differences being that: (1) we did not include two optional ingredients (insulin, antibiotics), and (2) we seeded at a higher density since the publication started with iPSC-derived neural stem cells instead of the expanded iPSC-derived NPCs as described below.

Briefly, assuming 0% loss upon reviving, we first thawed all cell lines in parallel NPCs (P.3) at an average density of 200-250,000 cells/cm2 onto PLO+laminin-coated plates in NPM containing 10uM Y-27632; the day of thaw, these cells were considered 25DPI. NPCs were expanded in NPM, with daily media changes, for ∼7-9 days (∼32-34 DPI) at which point they were reseeded at a density of ∼175,200 cells/cm2 in NPM containing 10uM Y-27632 (now P.4). After this reseeding, NPCs were expanded for another 6-8 days until 40DPI, at which time they were used to seed for neuron differentiation.

At 40DPI, or 15 days after thawing and passaging once, these iPSC-derived NPCs were harvested with accutase, counted, and reseeded at a density of ∼105,250 cells/cm2 in neuron differentiation media supplemented with Y-27632 (10uM, only included during initial seeding); the neuron differentiation media’s final composition is: 1X DMEM/F:12 (Gibco 11-320-033), 1X N-2 (Gibco 17-502-048), and 1X B-27 (Gibco 17-504-044). The day cells were seeded in this neuron media marked day 0 of neuron differentiation -- and cultures had complete media changes every two days – which for simplicity was scheduled for every Monday, Wednesday, and Friday when the differentiation was started on a Monday. This neuron differentiation continued until day 30, at which time neurons stained robustly for TUBB3 – yielding an ∼80% TUBB3-positive uniform population of neurons. Neuronal cultures generally did not need to be passaged during this 30-day protocol, but on the rare occasion that cultures became confluent or lifted – neurons were reseeded at the same density (∼105,250 cells/cm2) as previously described.

### 9. Quantitative IF: staining protocol and confocal imaging

All cell types were seeded into matrix-coated 8-chamber slides (Thermo Nunc Lab-Tek II Chamber Slide System 12-565-8) using their typical passaging protocol and upon reaching appropriate confluency were washed once with 1X cold DPBS (Gibco 14-190-144), followed by 10 minute room temperature fixation with cold 4% PFA (Thermofisher AAJ19943K2). After fixation, cells were washed once with cold 1X DPBS to remove excess fixative and either stored at 4C for later processing in 1X DPBS with parafilm to prevent evaporation or cells were directly processed for staining.

Processing for staining entailed the following sequential steps: permeabilization, blocking, incubation with primary antibody overnight at 4C, triple washes, incubation with secondary antibody for 1 hour at room temperature, triple washes, mounting with prolong glass hard-setting medium, ∼24 hour curing of prolong glass, and sealing cured slides with nail polish. Permeabilization was performed for 10 minutes at room temperature using the following room temperature solution: 1xDPBS + 5% donkey serum + 0.1% tritonX-100 (Sigma #X100-1L). The following room temperature blocking buffer was used both for blocking and to dilute antibodies: 1xDPBS + 5% donkey serum. Importantly, since all secondary antibodies used in this study were raised in donkey hosts, to reduce background we prepared these solutions using 100% donkey serum (60mg/mL in water, Jackson Lab 017-000-121).

Blocking was performed for 10 minutes at room temperature. All primary antibodies were added to room temperature blocking buffer using their respective dilutions (**Supplementary Table 5**). After 4C overnight primary antibody incubation elapsed, to remove excess antibody, slides were triple washed using 1X cold DPBS, with each wash consisting of a 5 minute incubation at room temperature. After triple washing, ProLong™ glass media containing Hoechst 33342 (invitrogen P36985) was brought to room temperature per manufacturer instructions. All secondary antibodies were added to room temperature blocking buffer using their respective dilutions. After 1 hour of room temperature incubation with secondary antibodies, slides were triple washed using the same process as before. Slides were then mounted with prolong glass using 50 x 24 mm and 0.16-0.19 mm glass coverslips (Fisher 12-541-055). Per manufacturer instructions, slides were allowed to cure for ∼24 hours at room temperature in the dark before sealing slides with OPI clear nail polish. Sealed slides were then immediately imaged on the confocal or stored at 4C in the dark for later processing. For each marker, 5-10 representative images were taken for each cell line across all cell types using a Zeiss LSM880 confocal imaging system with glycerol between 40x-63x objectives - resulting in a total range of representatives images per analysis for each group: 20-40 images for controls, 10-20 images for early-truncating, and 15-30 images for late-truncating lines.

### 10. Quantitative IF: confocal assessment of “mean per cell expression” as “mean fluorescent intensity per cell”

For all “mean fluorescent intensity per cell” results, image analysis was performed using Biodock (https://www.biodock.ai/), an artificial intelligence (AI)-assisted quantification platform, as previously described ^26–28^. Under “Project Setting,” the channel settings were rearranged to match the fluorescent channels of the images uploaded; channel 1 was matched to the green channel (i.e. protein of interest labeled by Alexa488), channel 2 was matched to the blue channel (i.e. nuclei labeled by Hoechst), and channel 3 was matched with the red channel (i.e. protein of interest labeled by Alexa555). For our biodock analyses, we used the “Object-Based” analysis mode, which is better at detecting objects/cells with defined areas.

Briefly, the following process was performed for each cell type -- resulting in four unique AI models, with one trained AI-model generated per cell type (iPSCs, NPCs at days 7 and 15, neurons); to train an accurate and precise AI-model for each cell type, the following was performed on random images from each cell line. First, all images (in .tif format) were uploaded to the Biodock website. Second, in biodock, to quantify the overall fluorescent intensity per image, using the Alexa488 and Alexa555 images, we defined the “whole image” label as the perimeter of the entire image. Third, to quantify the overall cell count per image, using the Hoechst channel images, we defined the “nuclei” label as the perimeter of each Hoechst-positive object. Each AI-model was iteratively trained and optimized until both objects, “whole image” and “nuclei”, were labeled with near perfect accuracy (>95%). Once trained, these four cell type-specific AI-models were then run on all images from their respective cell type using batch processing in biodock - after which all outputs were manually inspected to ensure artifact-free quantitative measurements; any mislabeled objects were corrected using the post-analysis editing feature on Biodock. Each biodock analysis produced a single .csv file that contained: the image filename, the object class assigned during labeling, and the integrated intensity of each channel per detected object (i.e. total fluorescent signal per object); these biodock .csv files were the inputs for the downstream statistical analysis in R as described below.

Downstream statistical analysis was performed in R using standard packages like tidyverse, ggpubr, and scales. Briefly, the following process was performed on each biodock .csv file. First, images whose filenames did not match the expected naming convention were excluded -- with the remaining filenames parsed to assign each image quantification to its corresponding identifiers that were encoded in the filename string: slide, well, cell line, marker (i.e. protein of interest stained), and KAT6A genotype group (i.e. Control, Early, or Late). Second, for each image, we defined the overall cell count as the total number of objects labeled as “nuclei” and the overall fluorescent signal was defined as the integrated intensity of the corresponding “whole image” object. Third, for each channel, the “mean fluorescent intensity per cell” was calculated by dividing the overall fluorescent intensity by the overall cell count of said image; defined in the previous sentence. Lastly, to determine whether KAT6A genotype influenced per cell expression of each marker (i.e. protein expression), we statistically compared the “mean fluorescent intensity per cell” of all images across the KAT6A genotype groups using unpaired two-sided t-tests, with a p < 0.05 set as the threshold for significance; here, individual images were the unit of analysis and p-values were corrected using Benjamini-Hochberg procedure. Results are plotted as box plots with individual images overlaid as points (**Fig. 2E,I**; **Fig. 5K; extended data figs. 2-3**) and the statistical analysis of each marker quantification is provided (**Supplementary Table 6**).

### 11. Quantitative IF: confocal assessment of “mean per cell morphology” as “mean cytoskeleton and nuclei size per cell” and “mean % TUBB3 positive”

To test if there was a significant difference in the size of day-70 neuronal culture cells across the three groups (controls, early-truncating, late-truncating), using IF and biodock, we quantified the mean area of TUBB3- and Hoechst-staining in individual cells to calculate the “mean cytoskeleton and nuclei size per cell”, respectively. Similar to the quantitative IF described above, this imaging analysis was performed using Biodock (https://www.biodock.ai/) ^26–28^. In this cell morphology analysis, we trained an additional AI-model to detect individual “cytoskeleton” and “nuclei” contours as objects instead of detecting “whole image” and “nuclei” objects as was done in the “mean fluorescent intensity per cell” analysis.

To train this new cell morphology model in biodock, two images from each cell line were randomly selected and used to define which objects should be labeled as “cytoskeleton” and “nuclei”. First, all of the TUBB3-Alexa555/Hoescht images (in .tif format) were uploaded to Biodock. Second, in biodock, to quantify the cytoskeleton size of individual neuronal cells, using the Alexa555 channel images, we defined the “cytoskeleton” label as the perimeter of each TUBB3-positive object. Third, in biodock, to quantify the nuclei size of individual neuronal cells, using the Hoechst channel images, we defined the “nuclei” label as the perimeter of each Hoechst-positive object. This AI-model was iteratively trained and optimized until both objects, “cytoskeleton” and “nuclei”, were labeled with ∼90% accuracy. Once trained, it was run on all images using batch processing in biodock - after which outputs were manually inspected to ensure artifact-free quantitative measurements; any mislabeled objects were corrected using the post-analysis editing feature on Biodock. This analysis produced a single .csv file containing the image filename, the object class assigned during labeling, and the area of each detected object in pixels (i.e. “neuron morphology output”); this file was the input for both downstream statistical analyses described below that were performed in R using standard packages like tidyverse, ggpubr, and scales.

For both downstream statistical analyses, using this “neuron morphology output” file, images whose filenames did not match the expected naming convention were excluded -- with the remaining filenames parsed to assign each image quantification to its corresponding identifiers that were encoded in the filename string: slide, well, cell line, and KAT6A genotype group (i.e. Control, Early, or Late).

To determine if KAT6A genotype influenced per cell morphology, using this “neuron morphology output” file, we statistically compared the following across the KAT6A genotype groups with unpaired two-sided t-tests: (1) areas (in pixels) of all “nuclei” objects and (2) areas (in pixels) of all “cytoskeleton” objects; a p-value < 0.05 was set as the threshold for significance; here, individual cells were the unit of analysis and p-values were corrected using Benjamini-Hochberg procedure. These results are plotted as violin plots with individual cells overlaid as points (**Fig. 2G-H**) and the statistical analysis of this quantification is provided (**Supplementary Table 7**).

To determine if KAT6A genotype influenced neuronal induction, using the same “neuron morphology output” file, we calculated the “% TUBB3 positive” per image by dividing the total number of “cytoskeleton” objects by the total number of “nuclei” objects then multiplied by 100. Then we statistically compared the “mean % TUBB3 positive” across the KAT6A genotype groups using unpaired two-sided t-tests with a p-value < 0.05 set as the threshold for significance; here, individual images were the unit of analysis and p-values were corrected using Benjamini-Hochberg procedure. These results are plotted as bar plots of the group mean ± SEM with individual images overlaid as points (**Fig. 2I**) and the statistical analysis of this quantification is provided (**Supplementary Table 7**).

### 12. Protein extraction and quantification

Whole cell lysate (WCL) was isolated on ice or at 4C from pelleted cells using 1X cell lysis buffer (Cell Signaling #9803) containing a 1X Protease and Phosphatase Inhibitor Cocktail (Thermo Scientific #78442) per manufacturer instructions. Prior to western blotting and proteomic mass spectrometry, the concentration of each protein extract was quantified in duplicate or triplicate using a Pierce BCA protein assay kit (Thermo Scientific #23225) and results were read on a Synergy H1 Microplate Reader using BioTek Gen5-3.04 software per manufacturer instructions. All protein extracts were handled on ice and stored at −80C.

### 13. Western blotting protocol

Protein extractions were freshly prepared under denaturing conditions by diluting samples in 4x Laemmli Sample Buffer (Biorad #1610747) supplemented with 14.3M β-mercaptoethanol (BME; Sigma #M-7522) per manufacturer instructions to obtain samples with a final concentration of 1X Laemmli Sample Buffer with ∼355 mM BME. Prepared samples were boiled for 10 minutes at 100°C in a PCR thermocycler, followed by loading 20ug per lane onto a 4–20% Stain-free Protein Gel (Biorad 5678094) with 1uL of Precision Plus Protein Dual Color Standards (Biorad 1610374). Samples were then run in gel electrophoresis system (Biorad 1656019) using 1X Tris-Glycine-SDS running buffer (Fisher Scientific BP13414) for 80 minutes at 130V.

Proteins were transferred from the gel onto nitrocellulose membranes (Biorad #1704271) using a semi-dry blotting transfer system (Biorad #1704150) and size-appropriate settings. Membranes were then blocked at room temperature for 1 hour with gentle agitation in a blocking buffer made of: 5% non-fat milk (Nestlé #12428935), 0.1% Tween20 (Fisher Scientific #BP337-100), and 1X Tris-Buffered Saline (TBS; Boston Bioproducts #BM3014L). After blocking, primary antibodies were diluted in blocking buffer and incubated with membranes at 4°C overnight with gentle agitation. The following day, membranes were washed 3 times at room temperature in 1X TBS supplemented with 0.1% Tween 20 (TBST), with gentle agitation for 10 minutes per wash, to remove excess antibodies. Membranes were then incubated for 1 hour at room temperature with secondary antibodies using gentle agitation in the dark; similar to primary antibody solutions, all secondary antibodies were diluted in blocking buffer. Next, membranes were washed 3 times at room temperature in TBST with gentle agitation for 10 minutes each wash to remove excess antibodies.

Lastly, the fluorescent signals from the resulting membranes was visualized on a ChemiDoc Imaging System with a black tray (Biorad #12003028) using multiplexed-fluorescent imaging settings. All western blot-related antibodies were prepared using their respective dilutions.

### 14. Western blotting quantification

To quantify the changes in relative protein abundance of each antigen of interest from the fluorescent western blots, we imported the blot images from each channel (i.e. IRdye680, IRdye800, or Rhodamine) into ImageJ ^29^. Once each image was imported into ImageJ, we selected each band of interest with the rectangular tool in ImageJ and ran a widely-used macros plugin to automatically quantify and correct the regions of interest (ROI) for background noise; for band quantification, ‘total’ is defined as the “integrated intensity within the ROI without background correction” and the ‘signal’ is defined as “the background corrected integrated intensity”. Briefly, we used the following setting with the Band/Peak Quantification macros: background width pixel set to 3, estimate background from top/bottom, and background estimation calculated from median. The macros’ algorithm then performs the same tasks as commercially available quantification software like Image Studio Lite from LI-COR Biosciences ^30^. Next, to determine the relative abundance of each antigen of interest, the resulting ‘signal’ for the antigen of interest was then normalized to the ‘signal’ for each respective loading control band on the same exact blot.

Statistical analysis was performed on loading-control normalized signal values in Graphpad PRISM software using a one-way ANOVA with Tukey’s multiple comparisons test to compare the means across the three genotypes, applying a significance threshold of p-adj< 0.05 in accordance with accepted statistical standards ^31,32^. The band quantifications and statistical analysis are provided for each blot (**Supplementary Tables 8-10**). We also provide the uncropped western blots in source data.

### 15. RNA: extraction, quality-control, quantification

For whole transcriptome RNA-sequencing analysis, total RNA was extracted from cultured cells in technical duplicate or triplicate after pelleting cells, snap-freezing on dry ice, and storing at −80C in 1.5 mL polypropylene tubes (Thermo Scientific #3451). Total RNA was extracted from snap-frozen pellets using a PureLink RNA extraction kit (Invitrogen #12183018A) in conjunction with an on-column PureLink DNase treatment (Invitrogen #12185-010). All DNA-free RNA was eluted in 30 uL of sterile nuclease-free water (Invitrogen #10977023) then stored at −80°C after aliquoting 2uL for downstream quality control.

RNA extractions were diluted prior to quality control by combining 3uL stock with 3uL of nuclease-free water for a dilution factor of 2. To determine the concentration, 1 uL of diluted RNA was quantified in duplicate using a broad-range Qubit fluorometer (Invitrogen #Q10211) and the average value was used. To assess the RNA Integrity (RIN) before library preparation, 2uL of diluted RNA was analyzed on an Agilent 4150 Tapestation System using High Sensitivity RNA Screen Tapes (Agilent #5067-5579) and ladder (Agilent #5067-5581) per manufacturer instructions. RNA extractions with a RIN value less than 8.2 were not used to prepare subsequent RNA-sequencing libraries.

### 16. RNA-seq: library preparation, quality control, quantification, sequencing

Total RNA was converted into DNA libraries using a TruSeq Stranded Total RNA Library Prep Gold kit (Illumina #20020599) and 96-well plates (Thermo Scientific #N8010560), which were sealed with microfilms (Thermo Scientific #4306311). All nucleic acids in this protocol were stored at −80°C and all the following bead-based clean-up steps were performed using the following reagents: AMPure XP beads (Beckman Coulter #A63881), 200 proof ethanol (Fisher Scientific #BP2818-500), sterile nuclease-free water (Invitrogen #10977023), and a 96-well magnetic stand (Invitrogen #AM10027).

First, to ensure all RNA samples were converted into RNA-sequencing libraries using identical inputs, all RNA samples were diluted to the same concentration in ELB buffer (Illumina #20020599). Next, to simultaneously fragmented and depleted samples of ribosomal RNA (rRNA), ∼300 ng of diluted RNA was mixed with 8.5 uL of EPH buffer (Illumina #20020599) and 1 uL of FastSelect rRNA-depletion enzyme (Qiagen #335377). To complete the fragmentation and deletion process, this 17 uL reaction was then placed on a T100 Thermal Cycler (Biorad #1861096) using the following program recommended by the Qiagen FastSelect handbook: 94°C for 8 minutes, 75°C for 2 minutes, 70°C for 2 minutes, 65°C for 2 minutes, 60°C for 2 minutes, 55°C for 2 minutes, 37°C for 2 minutes, 25°C for 2 minutes, and then hold at 4°C.

The depleted and fragmented RNA was then converted into cDNA using random hexamers following the library preparation workflow described in Illumina’s TruSeq Stranded Total RNA Library Reference Guide (Document #1000000040499-v00). Specifically, the 17 uL reaction from the previous step was directly used to ‘synthesize first strand cDNA’ by combining this reaction with 8 uL of a master mix containing a ratio of 1 uL of Superscript Reverse Transcriptase II (Invitrogen #18064-014) to 9 uL of FSA (Illumina #20020599). After cDNA synthesis, the reactions were cleaned up using AMPure XP beads and then adenylated at the 3’ ends according to Illumina’s TruSeq Stranded Total RNA Library Reference Guide.

Non-overlapping unique dual indexes (“IDT for illumina - Truseq RNA UD Indexes v2 (96 indexes, 96 samples) -- cat#20040871”) were then added to each reaction and ligated according to manufacturer instructions. Following adapter ligation and AMPure XP bead clean up, the DNA libraries were PCR amplified using primers specific to fragments containing adapter sequences per manufacturer’s instructions. Enriched DNA fragments were then cleaned up using AMPure XP beads and final DNA libraries eluted in 30 uL of RSB (Illumina #20020599). Before storing at −80C, DNA libraries were aliquoted for quality control and quantification by diluting 3 uL of library with 3 uL of nuclease-free sterile water.

Before pooling the resulting double-stranded libraries for next-generation sequencing, all diluted library aliquots were quantified in duplicate using a Qubit dsDNA HS fluorometer (Invitrogen #Q32854). Also library quality was assessed by analyzing diluted library on an Agilent 4150 Tapestation System using D1000 HS Screen Tapes (Agilent #5067-5584). To determine the average size of each library in base pairs (bp), this screen tape data was analyzed on TapeStation software to include all fragments ranging from 100bp to 850bp. Using the aforementioned library preparation protocol, each library ranged from 300bp to 350bp, with an average size of 320bp. Next this data was used to calculate the molarity of each original DNA library stock using the following formula:

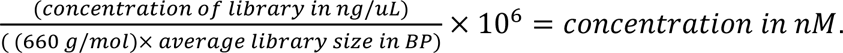

Lastly, RNA-seq libraries with non-overlapping barcodes (n=96 libraries) were diluted to 1.5 nM in sterile nuclease-free water and then equivalent volumes of each 1.5 nM library were combined to create a single equimolar pool. This 1.5 nM pool of libraries was re-analyzed using a Qubit dsDNA HS fluorometer and an Agilent 4150 Tapestation System to confirm correct dilution and pooling.

Then to reduce batch effect across our genomic assays, we sequenced our equimolar 1.5 nM RNA-seq pool with our equimolar 1.5 nM ATAC-seq pool on an illumina Novaseq6000 instrument in parallel on the same S4 flow cell, but separate lanes, using a 300-cycle kit with an XP-workflow and paired-end mode -- resulting in ∼150bp reads; we loaded 2 lanes of ATAC-seq and 2 lanes of RNA-seq libraries. The run was demultiplexed twice on illumina’s basespace server, once using RNA-seq adapter sequences to obtain RNA-seq FASTQ files and another time using ATAC-seq adapter sequences to obtain ATAC-seq FASTQ files as recommended by illumina. Adapter sequences were also removed on illumina’s basespace during the demultiplexing process -- resulting in adapter-free FASTQ files that were then downloaded and processed as described below.

### 17. RNA-seq bioinformatics pipeline

After next-generation sequencing, the RNA-seq data was demultiplexed, adapter sequences were removed, and fastq sequencing files were generated for all samples. Briefly, RNA-seq data was processed using the Arboleda lab’s RNA-seq pipeline ^33^ as follows. First, the quality of raw reads was assessed using FastQC (v0.11.8)^34^. Next, raw reads were mapped to the human genome by aligning raw reads against the Gencode human genome version hg38 (v31, GRCh38) ^35^ using STAR (v2.7.0e) to generate bam files ^36^. Gene expression was then estimated by generating gene count matrices from the STAR-mapped raw reads and a GTF file of the human gene annotations from Gencode’s primary assembly (v31, GRCh38) using FeatureCounts (v1.6.5)^37^ from the Subread package ^38^. In the gene count matrices, only reads that uniquely mapped to exons were counted for each gene.

BigWig coverage tracks were generated directly from the STAR-mapped BAM files output by the bioinformatics pipeline described above using bamCoverage from the deepTools suite ^39^. Coverage was calculated using the default bin size and normalized by counts per million mapped reads (CPM) to allow comparison of read density across samples. All STAR-mapped reads present in the BAM files were included. BigWig coverage tracks were visualized in Integrative Genomics Viewer (IGV) and annotated with UCSC’s genome browser using annotation tracks like ENCODE cis-Regulatory Elements and ENCODE Regulation.

### 18. RNA variant allele fraction analysis

Allele frequencies of truncated KAT6A variants from the ARTHS-derived lines were quantified from STAR-aligned, coordinate-sorted RNA-seq BAM files using samtools ^40^. For each RNA-seq sample, reads mapping to the KAT6A locus (chr8:41,929,479–42,051,994; GRCh38) were extracted with samtools view, and per-base pileups were generated with samtools mpileup against the GRCh38 reference genome (GENCODE v31). Bases with a phred quality score below 20 were excluded from the pileup. At each known truncated KAT6A variant position, the pileup output was parsed to count the number of reads carrying either the reference base and alternate bases. For each position, total read depth and alternate allele frequency (alternate read count / depth × 100) were recorded. Allele frequency results were then independently verified by manual inspection of truncated allele positions in IGV. Within each timepoint, we used two-sided Mann-Whitney U tests, a.k.a. Wilcoxon rank sum tests, to assess if there was a significant difference between the KAT6A RNA variant allele fractions of early- and late-truncating samples; this test was selected since the data was not normally distributed and the sample size was small; a p-value < 0.05 used as the significance threshold. The RNA variant allele fractions and summary statistics are provided (**Supplementary Tables 11**).

### 19. ATAC-seq: library preparation, quality control, sequencing

For each cell type (iPSC, day 15 NPCs, and neurons), ATAC-seq was performed with 50,000 cells in both control and patient-derived cell lines in technical triplicate. For each cell type, each line was lifted and counted in duplicate. ATAC-seq library generation was performed as previously described^41^. Briefly the cells were collected and treated with ATAC resuspension buffer for 3 minutes and the nuclei was pelleted at 500 RCF for 10 minutes at 4°C. The nuclei were then treated with Tn5 transposase (illumina #20034198) at 37°C for 25 minutes and processed for making libraries (Nextera XT DNA Library Preparation Kit (96 samples) # FC-131-1096). Samples were run on the Agilent DNA TapeStation to confirm tagmentation pattern.

Then non-overlapping genomically barcoded ATAC-seq libraries (n = 114 libraries) were all diluted to 1.5nM in sterile nuclease-free water and pooled at equimolar ratios to create a 1.5 nM pool. This 1.5 nM pool was re-quantified and re-tapestationed to ensure correct dilution and pooling as previously described. To reduce batch effect across our genomic assays, we sequenced our equimolar 1.5 nM RNA-seq and equimolar 1.5 nM ATAC-seq pools on an illumina Novaseq6000 instrument in parallel on the same S4 flow cell, but separate lanes, using a 300-cycle kit with an XP-workflow and paired-end modes -- resulting in ∼150bp reads; this processes is described above.

### 20. ATAC-seq bioinformatics pipeline

After next-generation sequencing, the ATAC-seq data was demultiplexed, adapter sequences were removed, and fastq sequencing files were generated for all samples. To ensure the precise identification of cell-type-specific changes in chromatin accessibility ^42^, all ATAC-seq fastq files were grouped by cell type upfront and then processed as follows using the Arboleda lab’s ATAC-seq pipeline ^33^. Similar to the Arboleda lab’s RNA-seq pipeline, first, the quality of raw reads was assessed using FastQC (v0.11.8) ^34^. Next, raw reads were mapped to the human genome by aligning raw reads against the Gencode human genome version hg38 (v31, GRCh38) ^35^ using BWA-MEM ^43,44^. BAM files were then sorted, indexed, and filtered with Samtools ^40^ to remove chrX, chrY, and MT reads. We then used Picard ^45^ to generate insert-size histograms and remove duplicates from BAM files; mapping statistics were also evaluated for BAM files with samtools. Next, we used MACS2 callpeak ^46,47^ to call narrow peaks for each sample; peaks that overlapped by at least 1 base were then merged using BEDtools merge ^48^. Next, reads overlapping these merged peaks were counted using featurecounts ^37^. We also annotated peaks using the “annotatePeaks.pl” command of the <u>H</u>ypergeometric <u>O</u>ptimization of <u>M</u>otif <u>E</u>n<u>R</u>ichment (HOMER, v4.9) package ^49,50^ -- allowing us to assign genomic features (e.g. exons, introns, promoters) to each peak.

BigWig coverage tracks were generated directly from the BWA-MEM-mapped BAM files output by the bioinformatics pipeline described above, which filtered out chrX, chrY, and mitochondrial reads. BigWig coverage tracks were generated using bamCoverage from the deepTools suite ^39^, with coverage calculated using the default bin size and normalized by counts per million mapped reads (CPM) to allow comparison of read density across samples. All reads present in these BWA-MEM-mapped BAM files were included. BigWig coverage tracks were visualized in Integrative Genomics Viewer (IGV) and annotated with UCSC’s genome browser using annotation tracks like ENCODE cis-Regulatory Elements and ENCODE Regulation.

### 21. DESeq2: differential expression and chromatin accessibility

Differential gene expression analysis was performed on the RNA-seq counts matrix using DESeq2 ^51,52^, which internally normalizes for library size and models read counts using a negative binomial distribution. Two comparisons were performed between the three groups (control, early-truncating, late-truncating): (1) early-truncating KAT6A versus control samples or (2) late-truncating KAT6A versus control samples; for both comparisons, sex was included as a covariate in the design formula (∼ sex + group) to adjust for sex since males and females were present in all three groups. While for the PF9363 treatment experiments, we compared treated versus untreated samples, adjusting for cell line by including it as a covariate (∼ cell_line + treatment) to isolate the effect of PF9363 across biological replicates. All log2 fold changes, log2FC, are reported as log₂(truncating/control) or log₂(treated/untreated). Hypothesis testing was performed with the Wald test and p-values were adjusted for multiple testing with Benjamini–Hochberg. Log2FC were shrunk using the lfcShrink() function with the apeglm estimator^53^ for approximate posterior estimation of GLM coefficients. Genes with an adjusted p-value < 0.05 and |log2FC| > 0.58 were considered as significantly differentially expressed genes (sigDEGs) and used in the downstream analyses as described in other sections.

Differentially accessible peaks were identified from the ATAC-seq peak count matrix using DESeq2, following the same statistical framework and comparison structures as the RNA-seq analysis described above; note modification: the DESeq2 analysis for the “day 15: NPC” ATAC-seq was adjusted for differentiation batch, so the design formula used for these specific comparisons was (∼ sex + batch + group). Significance was also assessed with the Wald test and p-values were also adjusted with Benjamini–Hochberg. Similarly, log2FC were shrunk using apeglm. We used the same significance cutoffs as was used to identify sigDEGs, peaks with an adjusted p-value < 0.05 and |log2FC| > 0.58 were considered significantly differentially accessible (sigPeaks) and used in the downstream analyses as described in other sections.

For all early- and late-truncating RNAseq and ATACseq comparisons from iPSCs, NPC day 15, and neurons -- we provide significant DESeq2 results as tables (**Supplementary Tables 12-23**). We also provide significant results for treatment-related RNAseq and ATACseq comparisons below in the treatment-related sections for clarity.

### 22. ClusterProfiler: enrichment analysis of RNA-seq & ATAC-seq results

We performed gene enrichment analyses using ClusterProfiler ^49^, with the Entrez gene IDs of genes classified as sigDEGs or sigPeaks by DESeq2 as inputs. The background gene universe was set to all the genes considered in the differential expression or differential accessibility analyses. We used the “compareCluster” function to run these gene enrichment analyses and used a corrected p-value < 0.05 as the threshold for significance (Benjamini–Hochberg adjusted). These enrichment analyses and visualization of results were performed according to the developer’s tutorials ^54,55^.

For all early- and late-truncating RNAseq and ATACseq comparisons from iPSCs, NPC day 15, and neurons -- we provide significant enrichment results as tables (**Supplementary Tables 24-26**). We also provide significant results for treatment-related RNAseq and ATACseq comparisons below in the treatment-related sections for clarity.

### 23. HOMER: motif analysis of RNA-seq and ATAC-seq results

Transcription factor binding motif analysis was performed on significant RNA-seq and ATAC-seq results described above using HOMER ^49,50^. For RNA-seq sigDEGs, enrichment was assessed in the promoters of each input gene list using HOMER’s “findMotifs.pl” function and the human genome (hg38) as reference; input text files were a single header-less column of Ensembl gene IDs. For ATAC-seq, enrichment was evaluated across the genomic intervals of sigPeaks using the “findMotifsGenome.pl” function and the human genome (hg38) as reference with a fixed window size of 200 bp that was centered at each sigPeak (-size 200); here, input BED files were six-column denoting: chromosome, start position, end position, a unique peak identifier, peak metadata, and strand information.

For the motif analysis of integrated RNAseq and ATACseq (RxA), we overlapped the gene names from RNAseq sigDEGs with gene names assigned to the ATACseq sigPeaks to identify genes that were dysregulated across both omics layers -- and then ran enrichment analysis using the associated BED files for those sigPeaks as previously described. For all motif analyses, HOMER’s default GC-matched background was used to identify enriched known and de novo motifs. Significantly enriched motifs were considered those with a p-value < 0.05. The HOMER motif enrichment results are provided as tables (**Supplementary Tables 27-38**).

### 24. ChIPseq and target gene lists

Only significant ChIPseq peak results were downloaded as BED files from ChIP-Atlas using their “Peak Browser” feature (https://chip-atlas.org/) for homo sapiens genome version hg38 ^56–59^. All downloads used a statistical significance threshold of Q-value < 1E-05 for peak calling and were calculated by the ChIP-Atlas using the MACS2 peak caller ^46^. To find high-confidence binding sites, significant peak data was downloaded from all available cell types. At the time of BED-file download, significant peak information for: KAT6A consisted of 9 independent datasets and FOS consisted of 70 independent datasets. KAT6A and FOS BED files were visualized (**Fig. 4**) with Integrative Genomics Viewer (IGV) software (ver 2.19.6) ^60–63^.

To construct the ChIPseq interaction network and target heatmaps (**Fig. 4**), human ChIPseq target gene lists (hg38) for the nodes in this network were downloaded from ChIP-Atlas ^56–59^ and TFLink gateway ^64^. The ChIPseq interactions between these nodes were visualized using Cytoscape ^65,66^ after excluding self-binding interactions for clarity. The non-ChIPseq RBFOX1 target list was constructed by downloading it from a study of RBP alternative-splicing events in RBP knockout mice RNAseq ^67^, converting mouse identifiers to human identifiers with biomaRt ^68,69^. We provide the target lists of KAT6A, FOS, PTBP1, and RBFOX1 used in this study (**Supplementary Tables 39-42**).

### 25. Proteomic mass spectrometry

Briefly, the proteomics mass spectrometry was conducted as described below in all nine biologically-independent iPSC lines used in this study and their neural derivatives (i.e. day 15 NPCs and neurons); these 9 cell lines were from 4 controls (00iCTR, 3iCTR, 83iCTR, 888iCTR), 2 early-truncating (1948iKAT6A, 1135iKAT6A), and 3 late-truncating (1618iKAT6A, 1521iKAT6A, 1105iKAT6A). Whole cell lysate was extracted from these samples and concentrations were measured as previously described in the ‘protein extraction and quantification’ methods section.

Normalized protein samples were sequentially reduced and alkylated using 5 mM tris(2-carboxyethyl)phosphine and 10 mM iodoacetamide, respectively followed by purification using the protein aggregation capture (PAC) approach ^70^, after which they were digested overnight at 37 °C with Lys-C and trypsin. The resulting peptides were subsequently dried and prepared for LC-MS/MS analysis.

The dried tryptic peptides were reconstituted in 5% formic acid and analyzed by liquid chromatography–mass spectrometry (LC-MS)-based proteomics using a Vanquish Neo UHPLC system coupled to an Orbitrap Astral mass spectrometer (Thermo Fisher Scientific, Bremen, Germany). In brief, peptides were loaded onto a PepSep C18 reverse-phase column (150 mm × 150 µm, 1.7 µm particle size) maintained at 59 °C and introduced into the Orbitrap Astral mass spectrometer through electrospray ionization. Chromatographic separation was carried out on the Vanquish Neo UHPLC using a trap-and-elute configuration. Mobile phase A consisted of water containing 0.1% formic acid, whereas mobile phase B contained acetonitrile with 0.1% formic acid. The 15-minute gradient was programmed as follows: 5% B from 0–1 min (flow rate = 2.45 µL/min), 5%–15% B from 1–5 min (1.75 µL/min), 15%–25% B from 5–12.6 min (1.75 µL/min), 25%–38% B from 12.6–13.6 min (1.75 µL/min), and 38%–80% B from 13.6–13.7 min (2.45 µL/min), followed by a hold at 80% B until 15 min (2.45 µL/min).

Data-independent acquisition (DIA) was carried out on the Astral mass spectrometer operating in positive electrospray ionization mode. MS1 spectra were recorded at a resolution of 240,000 over an m/z range of 380–980, with a normalized AGC target of 500% and a maximum injection time of 3 ms. DIA scans were performed using consecutive 4 m/z-wide isolation windows spanning the 380–980 m/z range. MS2 spectra were acquired at a resolution of 80,000 with a normalized higher-energy collisional dissociation (HCD) energy of 25%, a normalized AGC target of 500%, and a maximum injection time of 7 ms.

Thermo RAW files were processed using DIA-NN against an in silico spectral library generated from the Homo sapiens reference proteome (UP0000005640) ^71^. The DIA-NN output was subsequently evaluated in FragPipe Analyst to identify differentially regulated proteins using the limma package from R Bioconductor ^72^. A cutoff value of p-value < 0.05 was used to determine differentially enriched/abundant proteins in each of the following comparisons across 3 cell types (iPSCs, day 15 NPCs, and neurons): “Early-truncating vs. Control” and “Late-truncating vs. Control”; all log2(foldchange) results for these analyses are reported as log2(truncating/control). The significantly differentially abundant protein results from this proteomic mass spectrometry are included (**Supplementary Tables 43-48**).

### 26. Seahorse XF Assay: iPSC setup and settings

Two days prior to running the seahorse XF assay, iPSC lines were seeded as previously described into matrix-coated seahorse XF96 cell 96-well plates (Agilent 103794-100), avoiding the outer parameter of wells which were filled with 1X DPBS to prevent evaporation of inner wells. Briefly, vitronectin stock (0.5 mg/mL) was diluted 1/1000 as previously described then 106uL of diluted matrix was added to each well to obtain typical mass per cm2 (assumes 0.106 cm2) -- followed by overnight incubation at 37C before use. For all iPSC lines, ∼12 wells were plated at a total of ∼80,000 cells per well using accutase. The following lines were used and were at approximately passage 20 at the time of analysis: two controls (83iCTR, 888iCTR), one early-truncating (1135iKAT6A), and two late-truncating (1105iKAT6A, 1618iKAT6A). Cells were plated in 100 uL of complete E8 supplemented with 10 μM Y-27632, the following day a complete media change was performed to remove Y-27632 and allow cells to recover before analysis. Two days after seeding, the seahorse plate was submitted to the UCLA mitochondrial core for seahorse XF analysis. The core ran the plate using the following optimized settings for this cell type: 0.75 uM FCCP for injection 1, 1.35 uM FCCP for injection 2, 2 uM oligomycin, 1 uM Rotenone, and 2uM Antimycin A.

### 27. Seahorse XF Assay: NPC setup and settings

Four days prior to running the seahorse XF assay, NPC lines were seeded as previously described into matrix-coated seahorse XF96 cell 96-well plates (Agilent 103794-100), avoiding the outer parameter of wells which were filled with 1X DPBS to prevent evaporation of inner wells. Plates were double-coated with 106 uL of diluted PLO stock (stock: 5 mg/mL) and then 106 uL of diluted laminin stock (stock: 1 mg/mL) at 1/1000 as previously described to obtain typical mass per cm2 (assumes 0.106 cm2) and incubated overnight at 37C before use. For all NPC lines, ∼12 wells were plated at a total of ∼100,000 cells per well using accutase. The following lines were used and were at approximately passage 4 at the time of analysis: two controls (83iCTR, 888iCTR), one early-truncating (1135iKAT6A), and two late-truncating (1105iKAT6A, 1618iKAT6A). Cells were plated in 100 uL of complete neural progenitor expansion media (Stem Cell Technologies #05833) containing 10 μM Y-27632, the following day a complete media change was performed to remove Y-27632 and allow cells to recover before analysis. Four days after seeding, the seahorse plate was submitted to the UCLA mitochondrial core for seahorse XF analysis. The core ran the plate using the following optimized settings for this cell type: 0.75 uM FCCP for injection 1, 1.35 uM FCCP for injection 2, 2 uM oligomycin, 1 uM Rotenone, and 2uM Antimycin A.

### 28. Seahorse XF Assay: Neuron setup and settings

Two days prior to the end of the neuron differentiation protocol (28 of 30 days) and prior to running the seahorse XF assay, neuron lines were seeded as previously described into matrix-coated seahorse XF96 cell 96-well plates (Agilent 103794-100), avoiding the outer parameter of wells which were filled with 1X DPBS to prevent evaporation of inner wells. Plates were double-coated with 106 uL of diluted PLO stock (stock: 5 mg/mL) and then 106 uL of diluted laminin stock (stock: 1 mg/mL) at 1/1000 as previously described to obtain typical mass per cm2 (assumes 0.106 cm2) and incubated overnight at 37C before use. For all neuron lines, ∼5 wells were plated at a total of ∼16,000 cells per well using accutase. The following lines were used and were at day 30 of neuron differentiation at the time of analysis: four controls (83iCTR, 888iCTR, 00iCTR, 3iCTR), two early-truncating (1135iKAT6A, 1948iKAT6A), and two late-truncating (1521iKAT6A, 1618iKAT6A). Cells were plated in 100 uL of neuron media containing 10 μM Y-27632, the following day a complete media change was performed to remove Y-27632 and allow cells to recover before analysis. Two days after seeding, the seahorse plate was submitted to the UCLA mitochondrial core for seahorse XF analysis. The core ran the plate using the following optimized settings for this cell type: 1.25 uM FCCP for injection 1, 2.25 uM FCCP for injection 2, 2 uM oligomycin, 1 uM Rotenone, and 2uM Antimycin A.

### 29. Seahorse XF Assay: data analysis

Oxygen consumption rate (OCR) and the extracellular acidification rate (ECAR) were measured in a Seahorse XF96 Extracellular Flux Analyzer (Agilent Technologies). Cells were seeded and cultured on a Seahorse XF96 plate as previously described. On the day of the assay, cells were washed with Seahorse assay medium supplemented with 5 mM glucose, 2 mM glutamine, and 1 mM sodium pyruvate; pH 7.4. Compounds were injected sequentially during the assay resulting with oligomycin in Port A, FCCP in Ports B and C, and rotenone and antimycin in Port D. Rate measurements were normalized to cell number by counting Hoechst-stained nuclei (5 μg/mL Hoechst) on an Operetta High-Content Imaging System (PerkinElmer). Respirometry parameters were calculated as described by Divakaruni et al. and DeSousa et al. for the ATP production rates ^73,74^.

Statistical analysis was performed on cell count-normalized results in Graphpad PRISM software using either a one-way or two-way ANOVA, each with Tukey’s multiple comparisons test, to compare the means across the three genotypes, applying a significance threshold of p-adj < 0.05 in accordance with accepted statistical standards ^31,32^. The seahorse results and statistical analysis results for each cell type are provided (**Supplementary Tables 49-51**).

### 30. Quantitative IF of neuron mitochondria: “mean per cell GRP75 expression” and “mean per cell mitochondrial physiology”

*Confocal assessment of “mean per cell GRP75 expression” as “mean fluorescent intensity per cell” (****Fig. 5J-K***).

- **Seeding, staining, imaging**: Day-70 neuronal cultures were stained and imaged as previously described in “Quantitative IF: staining protocol and confocal imaging” section above. The following cell lines were used: 4 controls (83iCTR, 888iCTR, 00iCTR, 3iCTR), 2 early-truncating (1135iKAT6A, 1948iKAT6A), and 3 late-truncating (1521iKAT6A, 1618iKAT6A, 1105iKAT6A) -- and cultures were stained for GRP75 using a rabbit anti-GRP75 antibody (Cell Signaling #3593S).
- **Quantification and analysis**: To assess if there was a significant difference in GRP75 protein expression across the three groups (control, early, late) -- we quantified the “mean fluorescent intensity per cell” of GRP75 staining in day-70 neuronal cultures as exactly as described in the “Quantitative IF: confocal assessment of “mean per cell expression”” section above. These results are plotted as box plots with individual images overlaid as points (**Fig. 5K**) and the statistical analysis of this quantification is provided (**Supplementary Table 52**).

*High-content imaging assessment of “mean per cell mitochondrial physiology” as “mean mitochondrial content, morphology, and size per cell” (****Fig. 5L-N***).

- **Seeding**: Two days prior to the end of the neuron differentiation protocol, cells were seeded using accutase as previously described into matrix-coated 96-well black phenotyping plates (Revvity #6055302). Plates were double-coated with 200 uL of diluted PLO stock (stock: 5 mg/mL) and then 200 uL of diluted laminin stock (stock: 1 mg/mL) at 1/400 as previously described to obtain typical mass per cm2 (assumes 0.5 cm2) and prevent evaporation during overnight 37C incubation required prior to use. For all neuron lines, ∼5 wells were plated at a total of ∼16,000 cells per well. The following lines were used and were at day 30 of neuron differentiation at the time of analysis: four controls (83iCTR, 888iCTR, 00iCTR, 3iCTR), two early-truncating (1135iKAT6A, 1948iKAT6A), and two late-truncating (1521iKAT6A, 1618iKAT6A). One day after seeding, a complete media change was performed to remove Y-27632.
- **Staining**: Two days after seeding, which marked the end of the neuron differentiation protocol, day-70 cells were fixed with room temperature 4% PFA (Thermofisher AAJ19943K2) for 15 minutes at 37C to preserve mitochondrial morphology. After fixation, cells were washed four times with 1X DPBS to remove excess fixative. Cells were then stored in 1X DPBS and submitted to the UCLA mitochondrial core for processing. To assess mitochondrial morphology, the cells were stained, imagined, and analyzed as follows. Cells were permeabilized with 0.1% Triton X-100 (Sigma-Aldrich, T8787), 0.05% sodium deoxycholate (Sigma-Aldrich, 30970) in PBS, blocked, and stained with primary and secondary antibodies in blocking solution containing 5% donkey serum (Millipore Sigma, S30-M) using their respective dilutions; cultures were stained with mouse anti-GRP75 antibody (Antibody Inc NeuroMab, SKU: 75-127). After nuclear staining with DAPI, cells were kept in PBS at 4°C until imaging.
- **Imaging**: 3D image stacks were acquired at optimal Z-distance and reconstructed as Maximum Z-projections. Images were acquired with the Image Xpress Micro Confocal high-content imaging system (Molecular Devices) with a 40x/1.2 N.A. water objective.
- **Quantification and analysis**: A TopHat filter was applied to segment mitochondria, and a Gaussian Filter was applied to segment cellular area together with DAPI for nuclei detection. The images were then thresholded and transformed into a binary segmentation. To assess if there was a significant difference in mitochondrial physiology, we compared the segmented area of the following parameters across the groups (control, early, late) using unpaired two-sided t-tests: (1) “mean mitochondrial content per cell” as the “mean total mitochondrial area per cell”, (2) “mean mitochondrial morphology per cell” as the “mean shape factor per cell”, and (3) “mean mitochondrial size per cell” as the “mean mitochondrial area per cell”; individual wells were the unit of analysis and p-values were corrected for using Benjamini-Hochberg procedure. Results were plotted as violin plots with individual wells overlaid as points (**Fig. 5L-N**) and the statistical analysis of this quantification are provided (**Supplementary Table 52**).

### 31. Metabolite mass spectrometry: set up, input quantification, and extraction

To capture the metabolite profile of neurons at 30-days differentiation, cultures were reseeded at day 28 of 30 for metabolite mass spectrometry at a density of ∼52,631 cells per cm2 on PLO+Laminin-coated 12-well plates (Costar #3512) in 1000 uL of neuron media containing 10 μM Y-27632. The following day (day 29 of 30), Y-27632 was removed via complete media change. On day 30 of neuron differentiation, cultures were switched to labeled glucose (U-¹³C₆) media for ∼30 hours before metabolite extraction. The basal media used to prepare the labeling media contained no glucose, no glutamine, and no phenol red (Thermofisher A1443001). The final composition of the labeling media was: 2mM glutamine (Thermofisher 35050061), 1mM pyruvate (Thermofisher 11360070), 10mM U-¹³C₆ D-Glucose (Cambridge Isotope Laboratories CLM-1396-PK). After combining these elements, the labeling media was filter-sterilized.

For this analysis, each cell line was seeded in quadruplicate with three replicates used for extraction and one replicate used for counting to determine input amount; a minimum of 100,000 cells was used per condition. The following lines were used: four controls (00ictr, 3ictr, 83ictr, 888ictr), two early-truncating (1135ikat6ta, 1948ikat6a), and two late-truncating (1521ikat6a, 1618ikat6a). Each input sample was counted a minimum of four times with a countess II and the average of these values were later used for input-normalization of the resulting data. The remaining three wells were extracted on ice by first completely aspirating media off the wells, and then gently washing cells twice with ice cold 150 mM ammonium acetate (Sigma-Aldrich, A2706-100ML) without disturbing them. After completely aspirating off all ammonium acetate, metabolites were extracted by first adding −80C extraction solution (composed of 80% methanol supplemented with 100nM trifluoromethanesulfonate) to each well then placing the plate in the −80C freezer for 15 minutes. Afterwards, plates were placed back on ice and cells were scraped off (corning 3008) into the extraction solution and transferred into 2mL microcentrifuge tubes then vortexed vigorously. Next, tubes were centrifuged at 17,000g for 10 min at 4°C, then 250 uL of the supernatant was transferred to new 2mL tubes for evaporation. Metabolite extracts were immediately dried with N2 evaporation and stored at −80C for further processing as described below.

### 32. Metabolite mass spectrometry: data generation, processing, and analysis

Dried metabolites were reconstituted in 100 µL of a 50% acetonitrile (ACN) 50% dH20 solution. Samples were vortexed and spun down for 10 min at 17,000g. 70 µL of the supernatant was then transferred to HPLC glass vials. 10 µL of these metabolite solutions were injected per analysis. Samples were run on a Vanquish (Thermo Scientific) UHPLC system with mobile phase A (20mM ammonium carbonate, pH 9.7) and mobile phase B (100% ACN) at a flow rate of 150 µL/min on a SeQuant ZIC-pHILIC Polymeric column (2.1 × 150 mm 5 μm, EMD Millipore) at 35°C. Separation was achieved with a linear gradient from 20% A to 80% A in 20 min followed by a linear gradient from 80% A to 20% A from 20 min to 20.5 min. 20% A was then held from 20.5 min to 28 min. The UHPLC was coupled to a Q-Exactive (Thermo Scientific) mass analyzer running in polarity switching mode with spray-voltage=3.2kV, sheath-gas=40, aux-gas=15, sweep-gas=1, aux-gas-temp=350°C, and capillary-temp=275°C. For both polarities mass scan settings were kept at full-scan-range = (70-1000), ms1-resolution=70,000, max-injection-time=250ms, and AGC-target=1E6.

MS2 data was also collected from the top three most abundant singly-charged ions in each scan with normalized-collision-energy=35. Each of the resulting “.RAW” files was then centroided and converted into two “.mzXML” files (one for positive scans and one for negative scans) using msconvert from ProteoWizard ^75^. These “.mzXML” files were imported into the MZmine 2 software package ^76^. Ion chromatograms were generated from MS1 spectra via the built-in Automated Data Analysis Pipeline (ADAP) chromatogram module and peaks were detected via the ADAP wavelets algorithm ^77^.

Peaks were aligned across all samples via the Random sample consensus aligner module, gap-filled, and assigned identities using an exact mass MS1(+/-15ppm) and retention time RT (+/-0.5min) search of our in-house MS1-RT database. Peak boundaries and identifications were then further refined by manual curation. Peaks were quantified by area under the curve integration and exported as CSV files. If stable isotope tracing was used in the experiment, the peak areas were additionally processed via the R package AccuCor 2 to correct for natural isotope abundance ^78^. Peak areas for each sample were normalized by the measured area of the internal standard trifluoromethanesulfonate (present in the extraction buffer) and by the number of cells present in the extracted well.

We assessed global differences in overall pooled metabolite levels between the three groups (controls, early-truncating, and late-truncating) using input-normalized data using a one-way ANOVA with the following significance threshold cutoffs: * p-value < 0.05, ** p-value < 0.01, *** p-value < 0.001. This analysis identified 75 metabolites that were significantly differentially abundant across the three groups (**Supplementary Table 53**). The results of this ANOVA analysis are displayed as heatmaps of the log₂(FC) of total pooled metabolite levels in truncated and control neuronal cultures where log₂(FC) is calculated as log₂(individual sample value/average value of control group) and values are the sum of all its isotopomers after input normalization for cell number and trifluoromethanesulfonate (**Fig. 5, Extended Data Fig. 6**). We then performed a pathway-based metabolite set enrichment analysis on the compound names of these 75 significant metabolites with Metaboanalyst 6.0 (https://www.metaboanalyst.ca/) ^79^ using the following settings: Over Representation Analysis (ORA) algorithm, the SMPDB metabolite set library, and a threshold of FDR < 0.05 (**Supplementary Table 54**).

To determine specifically which groups were different and to what degree, we performed pair-wise comparisons between the three groups using input-normalized total pooled metabolite level (sum of all isotopomers) and the fractional contribution of the labeled nutrient (**Extended Data Fig. 6**) with unpaired t-tests using Graphpad PRISM software; the significance threshold was set to p-value < 0.05 for all pair-wise comparisons in accordance with accepted statistical standards appropriate for each comparison ^31,32^. We provide the input-normalized total pooled levels and fractional contribution values for all detected metabolites (**Supplementary Tables 55-56**).

### 33. Kainate treatment and IERG assessment

Neurons from one control line, one early-truncating line (1135iKAT6A), and one late-truncating line (1521iKAT6A) were seeded into 24-well plates with a density of ∼200,000 cells per well as previously described. The following day, cells were treated with kainate (Cayman Chemicals, Item No. 78050) for 1 hour. After treatment, cells were pelleted and RNA was extracted (Qiagen, Cat no. 74104) per manufacturer instructions. Extracted RNA was then submitted to UCLA’s Technology Center for Genomics & Bioinformatics (TCGB) core facility for RNA-seq library preparation, quality-control, sequencing, and data processing.

Briefly, RNA-seq libraries were prepared with ABclonal’s FAST mRNA Library Prep Kit, followed by sequencing on an illumina Novaseq X plus instrument using paired-end mode and a 100-cycle kit -- resulting in ∼50bp reads. Demultiplexed and adapter-free reads underwent quality control analysis and were aligned to the human genome (GRCh38) using STAR. Aligned reads were then quantified in Partek Flow against gene features defined by the Ensembl GTF annotation (v107) to generate a raw count matrix. Normalized RNA-seq counts were then generated by adding 1 × 10⁻⁴ to each raw count, followed by counts-per-million (CPM) normalization.

To determine the effect of kainate treatment on truncated lines, the normalized counts from early- and late-truncated RNA-seq samples were normalized to controls by dividing: (1) untreated truncated samples by untreated controls or (2) kainate-treated truncated samples by kainate-treated controls. We then calculated the log2 of each fold change (truncated/control), so gene expression increases and decreases were scaled symmetrically for statistical testing and visualization.

To assess the effects of kainate treatment across the two classes of truncating variants, statistical analysis was performed on the log₂(truncated/control) RNA-seq values by comparing untreated and kainate-treated samples using a RM one-way ANOVA (matched by gene) with the Geisser-Greenhouse correction and Bonferroni-corrected multiple comparisons in GraphPad Prism. This statistical analysis was applied to two gene sets, all protein-coding genes (n = 19,999) and high-confidence IERGs (n = 19) ^80^, to test the genotype-specific effect of kainate on global and activity-dependent gene expression effects, respectively. A significance threshold of p-adj < 0.05 was set, with per-gene log₂(truncated/control) RNAseq values and corresponding test statistics provided (**Supplementary Table 57**).

### 34. PF9363 treatments

Two control and three late-truncating NPC lines were treated with PF9363 at a final concentration of 1uM (Selleck #E0146) for different durations during the 30-day neuronal differentiation protocol; PF9363 was purchased at 10mM in DMSO, aliquoted on ice upon arrival then stored at −80C for future use. For maximum potency, PF9363 was thawed and diluted fresh every time the cultures needed their standard media change, with aliquots being thawed a maximum of two times before being discarded. On day 30 of neuron differentiation, cultures were harvested and processed for genomic analysis as described in the ATAC-seq and RNA-seq sections.

#### Control + PF9363

On day 0 of the 30-day neuronal differentiation protocol, two control NPC lines (00iCTR, 83iCTR) were seeded in neuron media containing 10uM Y-27632 at a density of ∼105,250 cells per cm2 on PLO+Laminin-coated plates and were cared for as previously described. In these control-related PF9363 experiments, four days after the two control NPC lines were seeded for neuronal differentiation (Day 4 of 30 days), cells were switched to neuron media containing freshly spiked-in 1uM PF9363 and received fresh PF9363-containing neuron media during every subsequent media change until the conclusion of the 30-day neuron differentiation protocol.

#### Late-truncating + PF9363

On day 0 of the 30-day neuronal differentiation protocol, three late-truncating NPC lines (1105iKAT6A, 1521iKAT6A, 1618iKAT6A) were seeded in neuron media containing 10uM Y-27632 at a density of ∼105,250 cells per cm2 on PLO+Laminin-coated plates and were cared for as previously detailed until PF9363 treatment. Then 11 days after their initial seeding (Day 11 of 30 days), late-truncating cells were overly confluent and an abnormal cellular morphology started to emerge, so cells were replated at the standard density of ∼105,250 cells per cm2. The following day, day 12 of 30 days, late-truncating cells were switched to neuron media supplemented with 1uM PF9363, receiving fresh PF9363-containing neuron media every media change until the conclusion of the 30-day differentiation protocol

Genomics analysis of both PF9363 paradigms: at the conclusion of PF9363 treatment, RNA-seq and ATAC-seq were performed and analyzed as described in the related sections above; we provide significant DESeq2 (**Supplementary Tables 58-60**) and clusterprofiler enrichment results (**Supplementary Tables 61-64**).

### 35. Factor-based SPEAR multi-omic integration

We fit SPEAR ^81^ on the RNA-seq, ATAC-seq, metabolite, and proteomic feature count matrices using two factors and default weighting, excluding the data from the KAT6A inhibitor experiment. The input counts were normalized using DESeq2 size factor normalization and variance stabilizing transform for the RNA-seq and ATAC-seq data, log transformation and z-score normalization for the metabolite data, and a log2 transformation with maximum intensity normalization for the proteomics data. We filtered to only include autosomal genes in the RNA-seq input to avoid sex-specific biases and then capped each modality to its 10000 most highly variable features.

After fitting, we identified factor 2 as the one affording separation between the late truncating cultures and the early truncating and control cultures. We used the resulting projection coefficients in the RNA-seq modality to rank genes based on their contribution to factor 2. To compare enrichment of Late: PF vs. DMSO differentially expressed genes from the inhibitor experiment in our gene ranking, we visualized the distribution of DE genes in the ranking based on SPEAR projection coefficients versus the ranking based on just the Late vs. Control DESeq2 log fold changes, showing enrichment for the former. We additionally tested for enrichment of Late: PF vs. DMSO differentially expressed genes in the top 500 genes nominated by the two rankings using a Chi-squared test, resulting in a p= 2.3E10^-5^. We provide the top 50 features from each modality (ATAC-seq, RNA-seq, proteomic and metabolite mass spectrometry) from Factor 2 (**Supplementary Table 65**).

### 36. Statistical analysis

Statistical testing was performed in R (version 4.1.1) and GraphPad Prism (version 11). Statistical tests were selected based on data type, sample size, and experimental setup -- with the specific statistical tests applied to each dataset explicitly stated in the relevant methods subsections above and individual figure legends.

## EXTENDED DATA FIGURES

**Extended Data Fig. 1:**
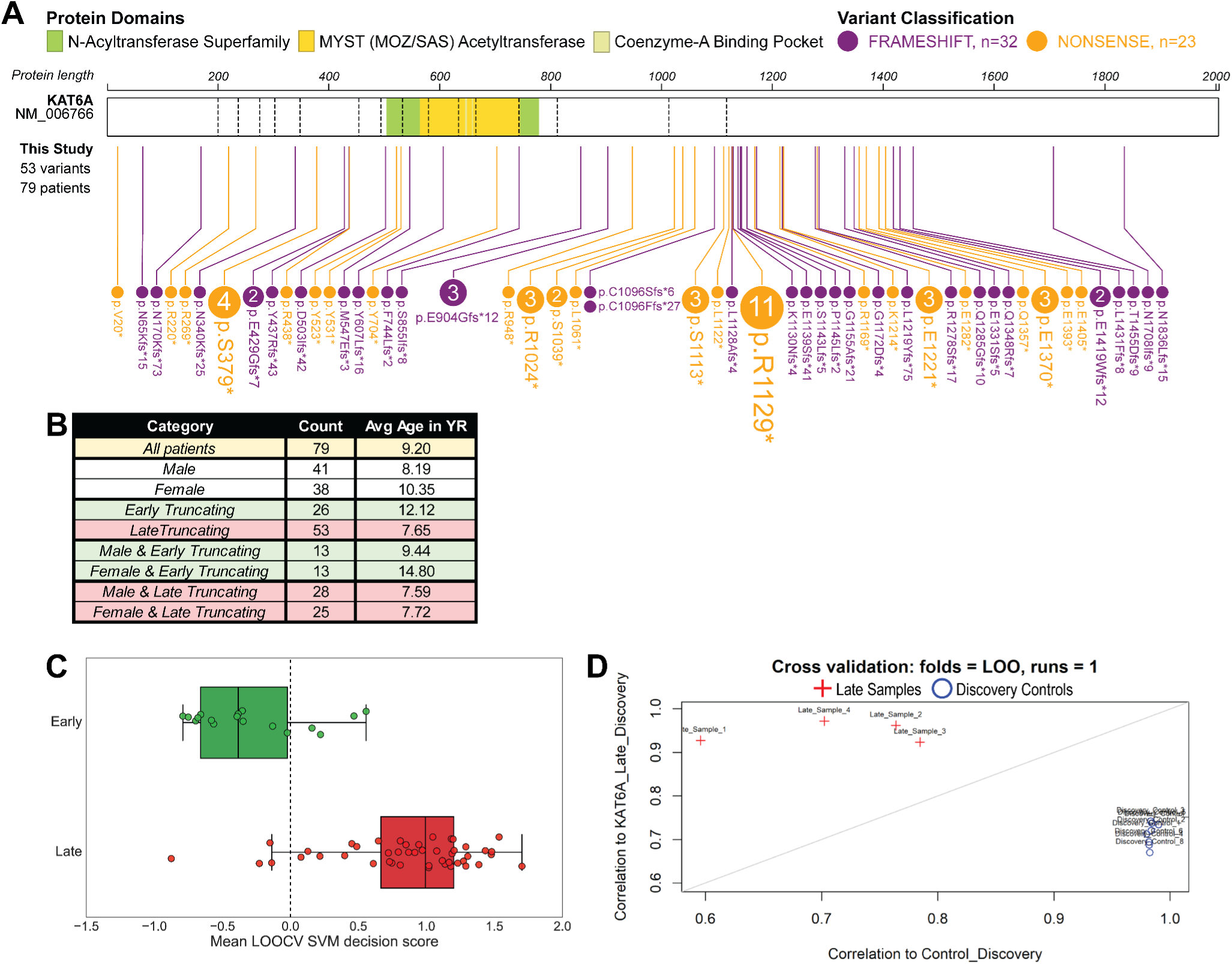
Variant landscape of all ARTHS cohorts and leave-one-out cross validation (LOOCV) of our GestaltMatcher and Blood DNAme Episignature analyses. **A**, Location of truncating *KAT6A* variants (n=53) across all ARTHS patients (n=79). **B**, Composition of 79 patients included across ARTHS cohorts: GestaltMatcher (n= 63 patients; 17 early, 46 late), Episignature (n=11 patients; 7 early, 4 late), and iPSCs (n=5 lines from 5 biologically-independent patients; 2 early, 3 late). **C**, GestaltMatcher facial analysis: image-level leave-one-out linear SVM classification; each point is one image from one patient; n= 63 patients; 17 early, 46 late. **D**, Blood DNAme Episignature analysis: Leave-one-out cross-validation analysis for the KAT6A late-truncating variant signature; each point is one individual patient DNAme sample; n= 11patients; 7 early, 4 late. Settings for each iteration of the LOOCV analysis were the same as those used in the initial generation of the signature (i.e. FDR corrected p-value < 0.05 and a |Δβ| > 0.1). Discovery samples are represented by red crosses, while discovery controls are represented by blue circles.

**Extended Data Fig. 2:**
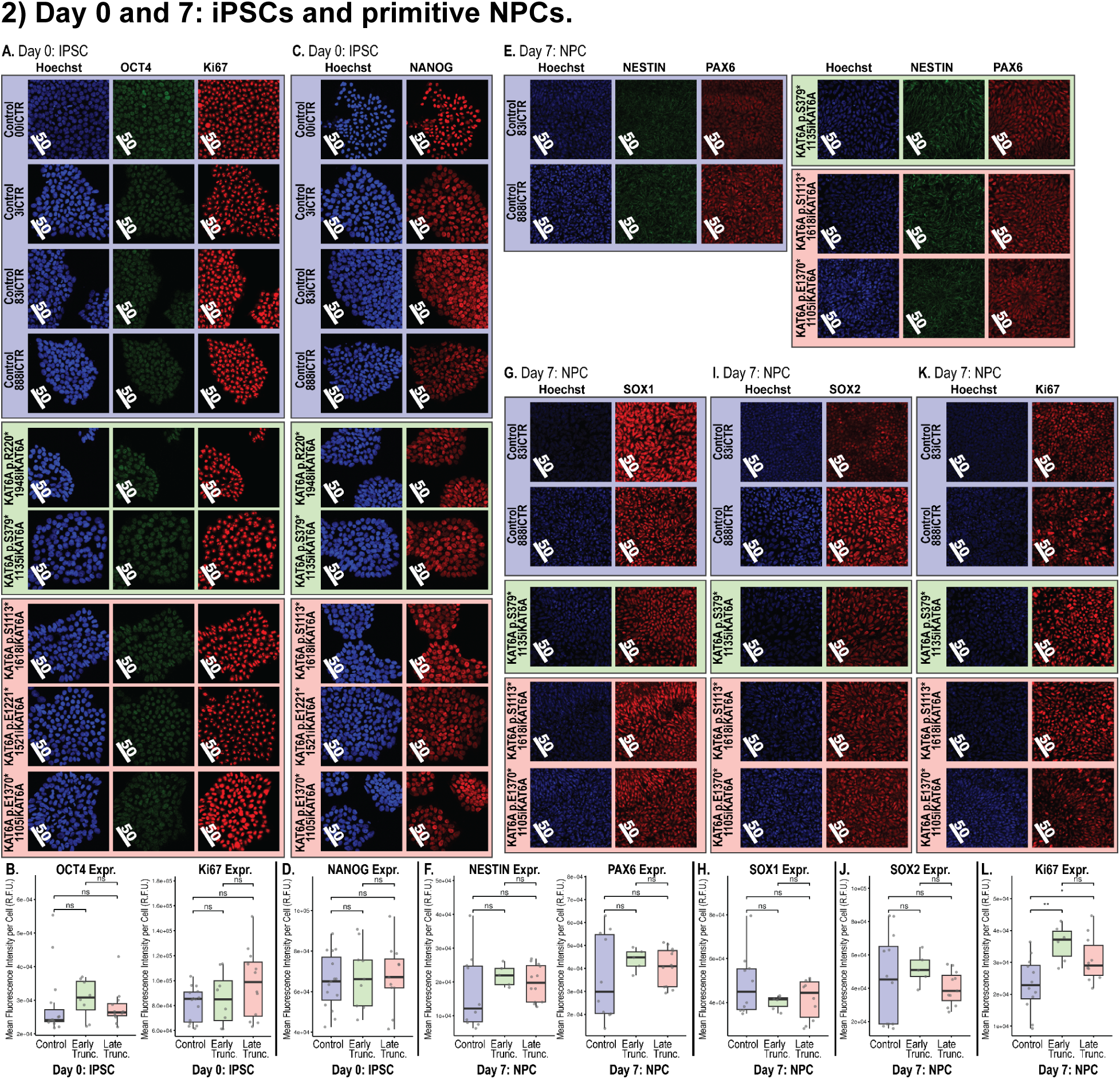
Assessment of starting (day 0: iPSC) stemness and defects at the inception of neural progenitor cell differentiation (day 7: NPC). **A-L**, Quantitative IF: comparing mean per cell fluorescent intensity (i.e. per cell protein expression) between 3 groups (control, early-truncating, late-truncating) using unpaired t-tests followed by BH multiple-testing correction; asterisks = p-adj < 0.05. Scale bars are shown in white and are all 50 um. All images are representative. **A-B,** Quantitative IF: day 0 IPSC cultures stained for OCT4 and Ki67. Box plot of mean per cell expression of markers shown in panel A. OCT4 (CTRL vs Early, p-adj= 0.501; CTRL vs Late, p-adj = 0.867) and Ki67 (CTRL vs Early, p-adj= 0.622; CTRL vs Late, p-adj = 0.077) **C-D,** Quantitative IF: day 0 IPSC cultures stained for NANOG. Box plot of mean per cell expression of marker shown in panel C. NANOG (CTRL vs Early, p-adj= 0.669670209; CTRL vs Late, p-adj = 0.661344446) **E-F,** Quantitative IF: day 7 NPC cultures stained for NESTIN and PAX6. Box plot of mean per cell expression of markers shown in panel E. NESTIN (CTRL vs Early, p-adj= 0.222; CTRL vs Late, p-adj = 0.496) and PAX6 (CTRL vs Early, p-adj= 0.256; CTRL vs Late, p-adj = 0.488). **G-H,** Quantitative IF: day 7 NPC cultures stained for SOX1. Box plot of mean per cell expression of marker shown in panel G. SOX1 (CTRL vs Early, p-adj= 0.104; CTRL vs Late, p-adj = 0.216) **I-J,** Quantitative IF: day 7 NPC cultures stained for SOX2. Box plot of mean per cell expression of marker shown in panel I. SOX2 (CTRL vs Early, p-adj= 0.479; CTRL vs Late, p-adj = 0.557) **K-L,** Quantitative IF: day 7 NPC cultures stained for Ki67. Box plot of mean per cell expression of marker shown in panel L. Ki67 (CTRL vs Early, p-adj= 0.002; CTRL vs Late, p-adj = 0.029)

**Extended Data Fig. 3:**
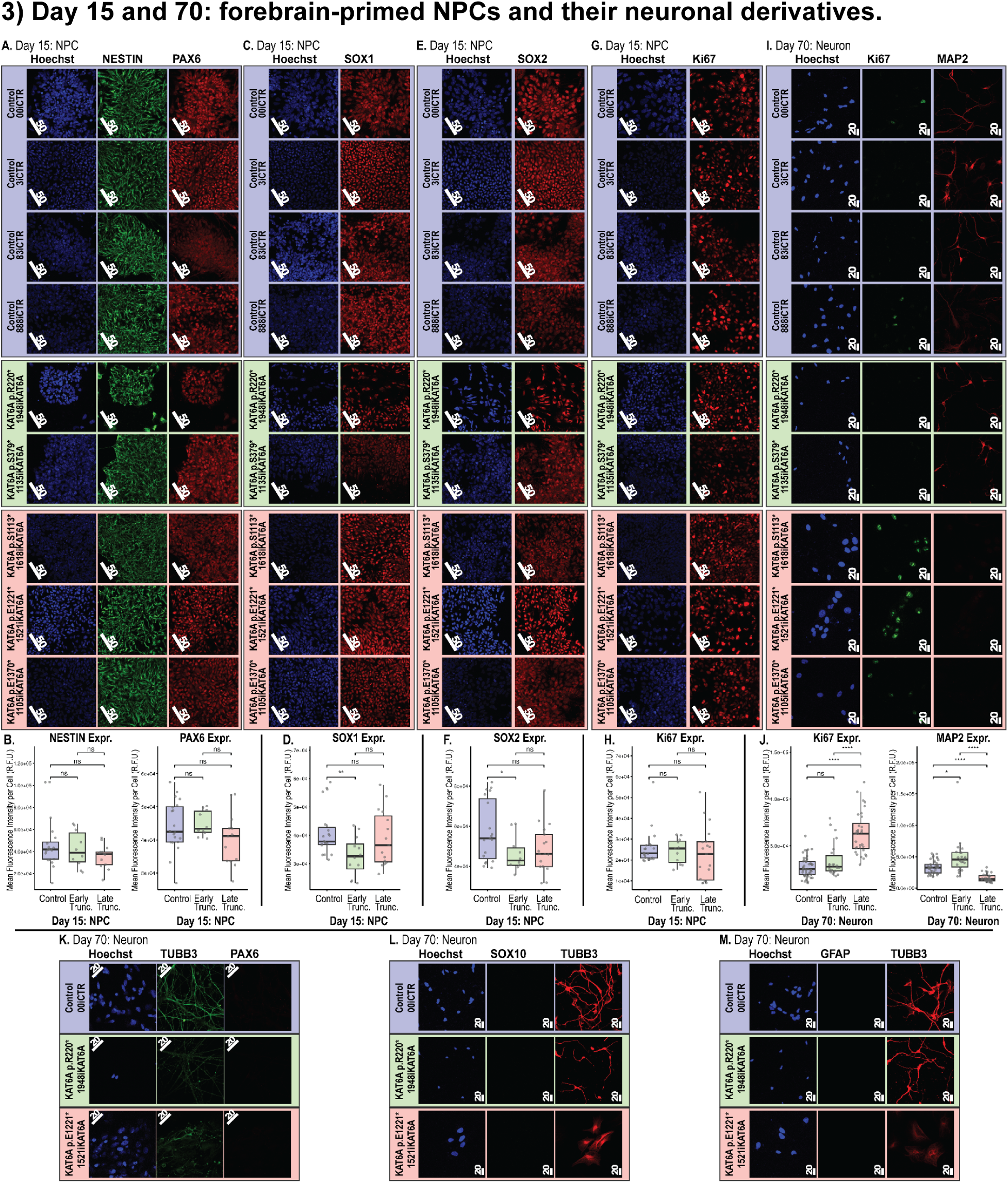
Assessment of NPC identity (day 15: NPCs) and aberrant neuronal differentiation (day 70: neuron). **A-J**, Quantitative IF: comparing mean per cell fluorescent intensity (i.e. per cell protein expression) between 3 groups (control, early-truncating, late-truncating) using unpaired t-tests followed by BH multiple-testing correction; asterisks = p-adj<0.05. Scale bars are shown in white, ranging from 20 um to 50 um. All images are representative. **A-B,** Quantitative IF: day 15 NPC cultures stained for NESTIN and PAX6. Box plot of mean per cell expression of markers shown in panel A. NESTIN (CTRL vs Early, p-adj=0.539; CTRL vs Late, p-adj = 0.072), PAX6 (CTRL vs Early, p-adj= 0.716; CTRL vs Late, p-adj = 0.170). **C-D,** Quantitative IF: day 15 NPC cultures stained for SOX1. Box plot of mean per cell expression of marker shown in panel C. SOX1 (CTRL vs Early, p-adj= 0.005; CTRL vs Late, p-adj = 0.471). **E-F,** Quantitative IF: day 15 NPC cultures stained for SOX2. Box plot of mean per cell expression of marker shown in panel E. SOX2 (CTRL vs Early, p-adj= 0.017; CTRL vs Late, p-adj = 0.063). **G-H,** Quantitative IF: day 15 NPC cultures stained for Ki67. Box plot of mean per cell expression of marker shown in panel G. Ki67 (CTRL vs Early, p-adj= 0.585; CTRL vs Late, p-adj = 0.404). **I-J,** Quantitative IF: day 70 neuronal cultures stained for Ki67 and MAP2. Box plot of mean per cell expression of markerS shown in panel I. Ki67 (CTRL vs Early, p-adj= 0.004; CTRL vs Late, p-adj = X); MAP2 (CTRL vs Early, p-adj= X; CTRL vs Late, p-adj = 4.2E-8). **K-M**, Representative IF images of day 70 neuron cultures from control, early-truncating, and late-truncating lines. No quantification was performed as cultures did not stain positive for non-neuronal markers (e.g. PAX6, SOX10, and GFAP). **K**, Representative IF images of day 70 neuronal cultures stained TUBB3 and PAX6. **L,** Representative IF images of day 70 neuronal cultures stained SOX10 and TUBB3. **M,** Representative IF images of day 70 neuronal cultures stained GFAP and TUBB3.

**Extended Data Fig. 4:**
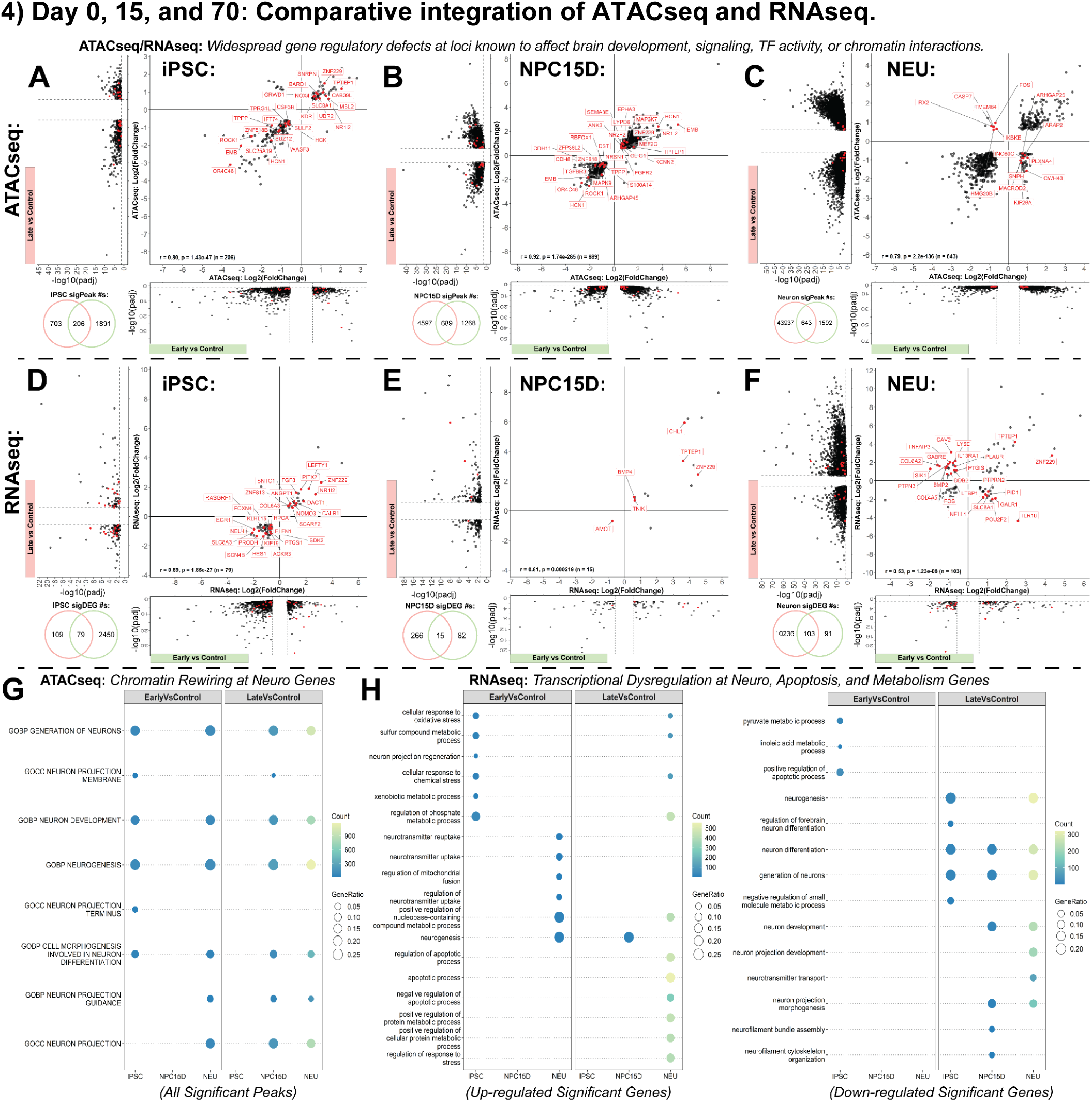
Gene regulatory rewiring: truncating KAT6A variants cause dysregulation across loci critical to proper brain development. **A-C**, Differential chromatin accessibility across three cell types: iPSCs, NPCs at day 15 (NPC15D), and neuronal cultures. All panels show: left volcano plot and Y-axis of center scatter plot display all significantly differentially accessible peaks (sigPeaks) in the comparison of late truncating vs control samples; bottom volcano plot and X-axis of center scatter plot display all sigPeaks in the comparison of early truncating vs control samples. Gene names and dots shown in red correspond to loci that affect brain development, signaling, TF activity or chromatin interactions. Pearson’s correlation was calculated between X and Y values in center scatter (printed in bottom lezft). **D-F**, Differential gene expression across three cell types: iPSCs, NPCs at day 15 (NPC15D), and neuronal cultures. All panels show: left volcano plot and Y-axis of center scatter plot display all significantly differentially expressed genes (sigDEGs) in the comparison of late truncating vs control samples; bottom volcano plot and X-axis of center scatter plot display all sigDEGs in the comparison of early truncating vs control samples. Gene names and dots shown in red correspond to loci that affect brain development, signaling, TF activity or chromatin interactions. Pearson’s correlation was calculated between X and Y values in center scatter (printed in bottom left). **G-H**, Over representation analysis of the significantly differentially regulated genes (i.e. sigDEGs, sigPeaks) across neuronal differentiation using an enrichment cutoff of padj<0.05. All log2(FC) are log2(truncating/control), with significant hits from RNAseq and ATACseq defined as genes with p-adjusted < 0.5 and an absolute value of log2(FC) > 0.58.

**Extended Data Fig. 5:**
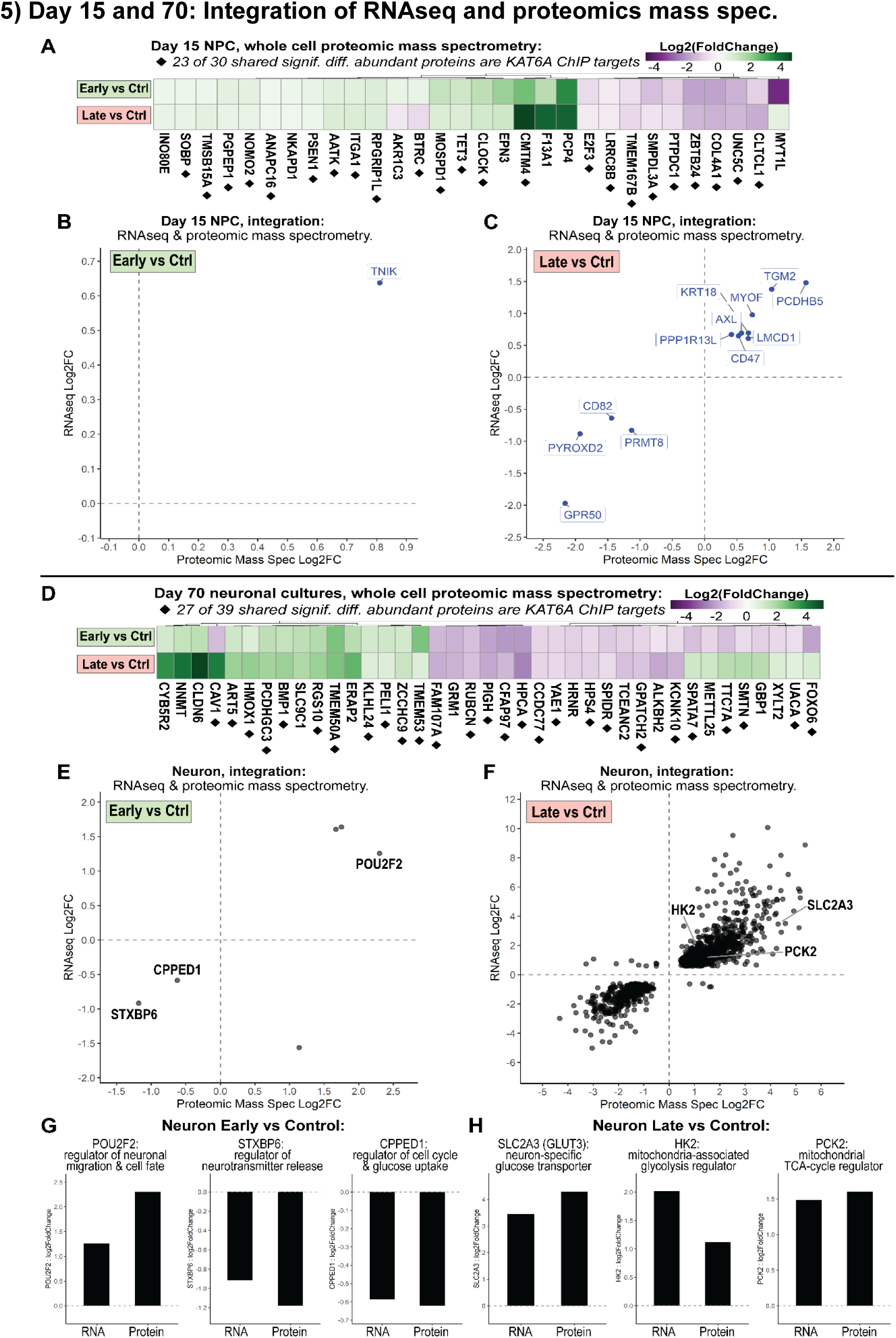
Truncating KAT6A variants cause proteome-level dysregulation of KAT6A ChIP targets related to metabolism and brain health genes. **A**, Protein mass spectrometry identifies 30 proteins to be significantly differentially abundant in the whole cell lysate of day 15 NPCs from both early- and late- truncating KAT6A samples compared to controls; 23 of 30 shared hits (76.7%) are direct KAT6A ChIP targets (◆ gene names) and 12 of 23 these shared targets (52.2%) affect metabolism (SMPDL3A), brain health (UNC5C, ZBTB24, E2F3, CLOCK, BTRC, SOBP, RPGRIP1L, COL4A1), or both (CLTCL1, TET3, PSEN1). **B-C**, Integration of significant results: RNAseq (y-axis) and proteomic mass spectrometry (x-axis) analysis of day 15 NPCs from early- and late-truncating lines identifies dysregulation of neuro- and metabolism related genes in cells harboring either class of KAT6A variant compared to controls; TNIK, a WNT pathway activator known to regulate neurodevelopment and metabolism, is significantly increased in early NPCs; while late NPCs exhibit significant misexpression of several genes related to neurodevelopment (e.g. AXL, PCDHB5) or metabolism (e.g. PYROXD2) or both (e.g. GPR50, PRMT8). **D**, Protein mass spectrometry identifies 39 proteins to be significantly differentially abundant in the whole cell lysate of neuron cultures from both early- and late-truncating KAT6A samples compared to controls; 27 of 39 shared hits (69.2%) are direct KAT6A ChIP targets (◆ gene names) and 18 of 27 shared targets (66.7%) have known roles in metabolism (KLHL24, BMP1, ART5), brain health (UACA, SPIDR, ZCCHC9, PCDHGC3), or both (FOXO6, KCNK10, HPS4, HPCA, PIGH, RUBCN, FAM107A, PELI1, RGS10, HMOX1, CAV1). **E-F**, Integration of significant results: RNAseq (y-axis) and proteomic mass spectrometry (x-axis) analysis of neuronal cultures from early- and late-truncating lines identifies dysregulation of neuro- and metabolism related genes in cells harboring either class of KAT6A variant compared to controls; labeled genes represent key examples. **G-H**, Gene expression barplots for key examples of significantly dysregulated neuro- and metabolism related genes in cells harboring either class of KAT6A variant compared to controls from panels E-F. All log2(FC) are log2(truncating/control), with significant hits from RNAseq and proteomics mass spectrometry defined as genes with either p-adjusted < 0.5 and an absolute value of log2(FC) > 0.58 or p-value<0.05, respectively.

**Extended Data Fig. 6:**
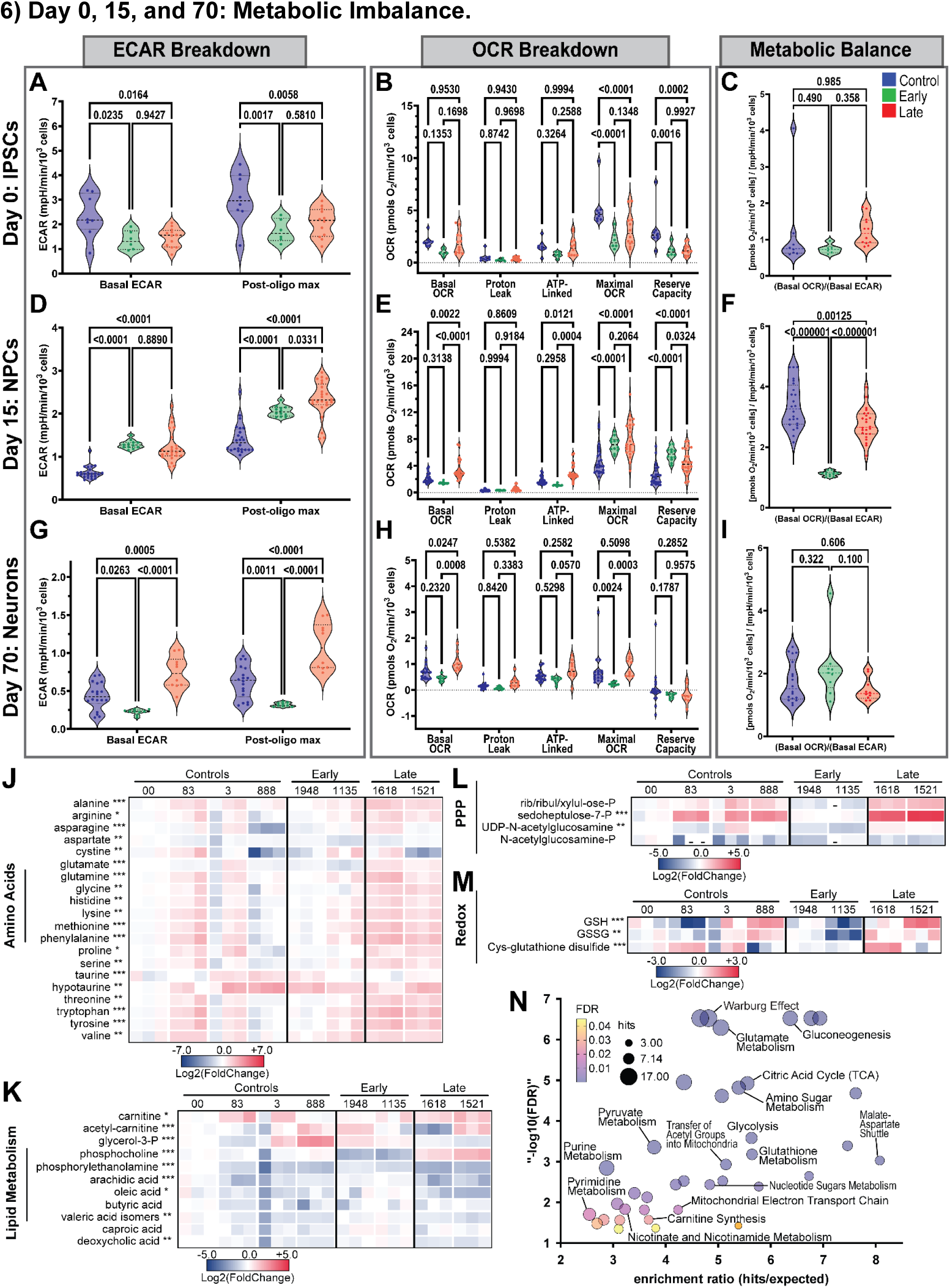
Early- and late-truncating KAT6A variants cause neuron-specific hypo- and hyper-metabolic states, respectively. **A-I**, Per cell mitochondrial bioenergetic profiles of iPSC, NPC, and neuronal cultures. All seahorse plots mean ± SEM and significant differences between three groups were assessed via two-way ANOVA (“ECAR breakdown” and “OCR breakdown” panels) and one-way ANOVA (“Metabolic Balance” panels) -- with both followed by Tukey’s post-hoc multiple comparison test (p-adj<0.05). **J-M**, Results of labeled glucose (U-¹³C₆) metabolite mass spectrometry: heatmaps of the log2(foldchange) of total pooled metabolite levels in truncated and control neuronal cultures; log2(FoldChange) is calculated as log2(individual sample value/average value of control group) where values are the sum of all its isotopomers after input normalization for cell number and trifluoromethanesulfonate. Heatmaps contain 34 of 75 differentially abundant metabolites: 20 amino acids, 9 lipid-related, 2 pentose phosphate pathway (PPP)-related, 3 redox-related; 12 of 75 significantly differentially abundant metabolites not included in heatmaps due to space constraints: sorbitol, n-acetylglutamate, betaine, cystathionine, folate, carnitine, nicotinamide, tryptophan, histidine, UDP-glucuronic acid, xylulose, s-(1;2-Dicarboxyethyl)glutathione. All significance tests were performed on normalized data with a one-way ANOVA; * p-value < 0.05, ** p-value < 0.01, *** p-value < 0.001. **N**, Results of D-Glucose (U-¹³C₆, 99%) metabolite mass spectrometry: 40 enriched metabolism terms (FDR < 0.05) generated by analyzing the list of 75 significantly differentially abundant metabolites from the one-way ANOVA analysis of total pooled metabolite levels (input-normalized) detected across control, early, and late neuronal cultures; this metabolite set enrichment analysis (MSEA) was performed via MetaboAnalyst’s ORA feature.

**Extended Data Fig. 7:**
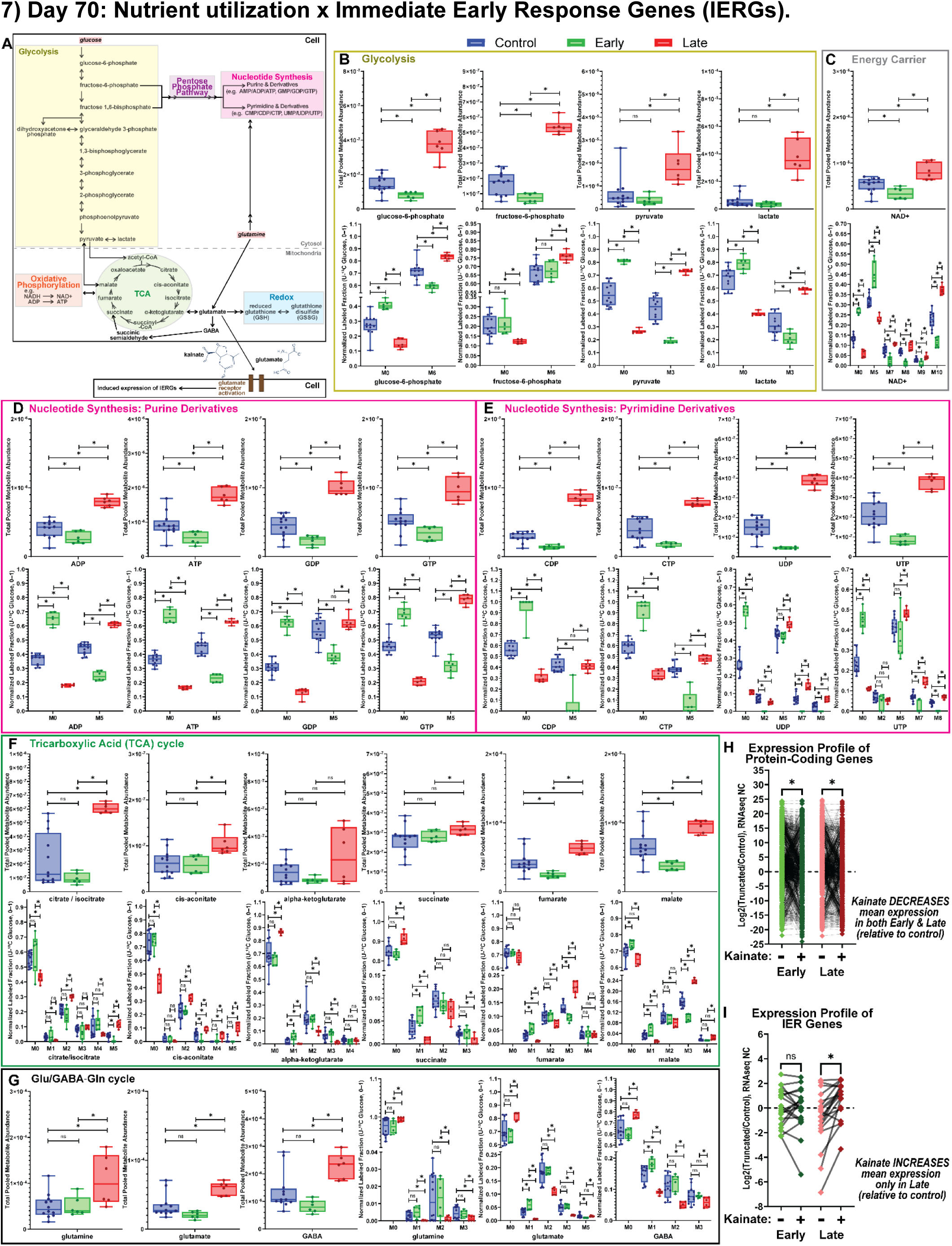
Early- and late-variants drive reduced and elevated glucose utilization/IERG expression, respectively, except late’s excess Glu/GABA-Gln metabolites are not glucose-derived. **A**, Diagram of core cellular metabolism pathways which processes nutrients (i.e. glucose, glutamine) and their known relation to inducing the expression of immediate early response genes (IERGs) through glutamate receptor activation mediated by glutamate or kainate, a potent neuroexcitatory compound that acts as an agonist for ionotropic glutamate receptors (AMPA and kainate types). **B-G**, Results of D-Glucose (U-¹³C₆, 99%) metabolite mass spectrometry performed on neuronal cultures from controls, early, late lines. For each metabolite, two normalized values are plotted across the 3 groups: the total pooled metabolite level (sum of all isotopomers) and the fractional contribution of the labeled nutrient (U-¹³C₆-Glucose) to its main isotopologues (sum of all M# is 1). All significance tests were performed on normalized data with unpaired t-test; * p-value < 0.05. **G**, Relative to controls and early truncating neuronal cultures, late truncating lines have excess glutamate (Glu), glutamine (Gln), and gamma-aminobutyric acid (GABA) that are not glucose-derived. **H-I**, Before & after line plots displaying normalized RNAseq expression profiles of all protein-coding genes or immediate early response (IER) genes from untreated (-) or kainate-treated (+) neuronal cultures; each points is the log2(foldchange) expression of individual genes within each gene set, where fold change values are the normalized counts of truncated samples divided by control samples. Statistical analysis was performed by comparing untreated and kainate-treated samples using a RM one-way ANOVA (matched by gene) with the Geisser-Greenhouse correction and Bonferroni-corrected multiple comparisons; asterisks = comparisons with p-adj<0.05. **H**, Expression profile of all protein-coding genes (n=19,999); there is a significant decrease in protein-coding gene expression for both classes of truncated samples upon treatment relative to controls, indicating equivalent global transcriptional responses to kainate treatment across genotypes. **I**, Expression profile of high-confidence IERGs (n=19); kainate treatment significantly and selectively induces IERG expression in late neurons, supporting that late’s excess Glu/GABA-Gln metabolites are functionally active as indicated by significant increase in IERG expression only in late samples.

**Extended Data Fig. 8:**
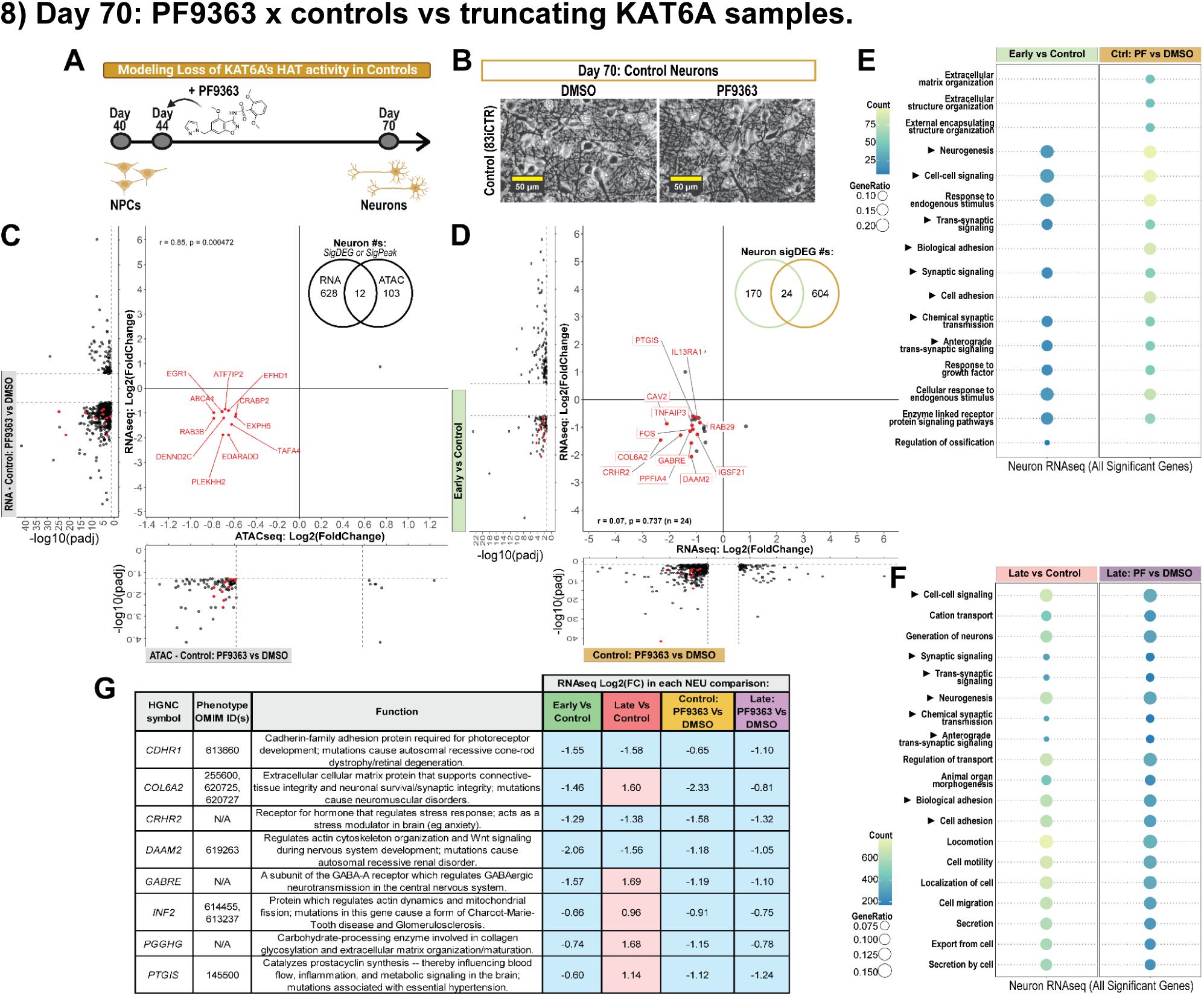
Effects of inhibiting KAT6A function in normotypic controls and its overlap with transcriptomic defects identified in truncating KAT6A samples. **A**, Overview of experimental design used to model loss of KAT6A’s histone acetyltransferase function during neuronal differentiation using PF9363 on controls; at day 4 of the 30-day neuron differentiation, or day 44 *in vitro*, PF9363 was added until the end of the protocol. **B**, Representative bright-field images of control neuronal cultures at day 70 *in vitro* after being treated with PF9363 as shown in panel a; scale bar is 50 µm. **C**, Direct gene regulatory effect of losing KAT6A’s histone acetyltransferase function during neuronal differentiation identified through incorporation of significantly differentially regulated genes across ATACseq (X-axis) and RNAseq (Y-axis) from experiments shown in panel a-b. **D**, Integration of neuron RNAseq: significantly differentially expressed genes (sigDEGs) from Early vs Control (left volcano) were overlapped with those from Control: PF9363 vs DMSO (bottom volcano). Center scatter plot and overlaid venn diagram identify 24 sigDEGs found in both RNAseq comparisons; genes labeled in red are known to affect brain development, signaling, transcription factor activity, or chromatin interactions. **E-F**, Overrepresentation pathway analysis of all significantly differentially expressed genes identified in each of the four neuron RNAseq comparisons presented in this study: “early vs control”, “late vs control”, “controls: PF9363 vs DMSO”, and “late truncating: PF9363 vs DMSO”. Eight pathways (marked with ▶) are consistently enriched amongst sigDEGs from all four comparisons shown. **G**, Known function of 8 genes (CDHR1, COL6A2, CRHR2, DAAM2, GABRE, INF2, PGGHG, PTGIS) that were significantly differentially expressed across all four neuron RNAseq comparisons – where the log2(foldchange) each “X” vs “Y” comparison is always log2(“X”/“Y”).

**Extended Data Fig. 9:**
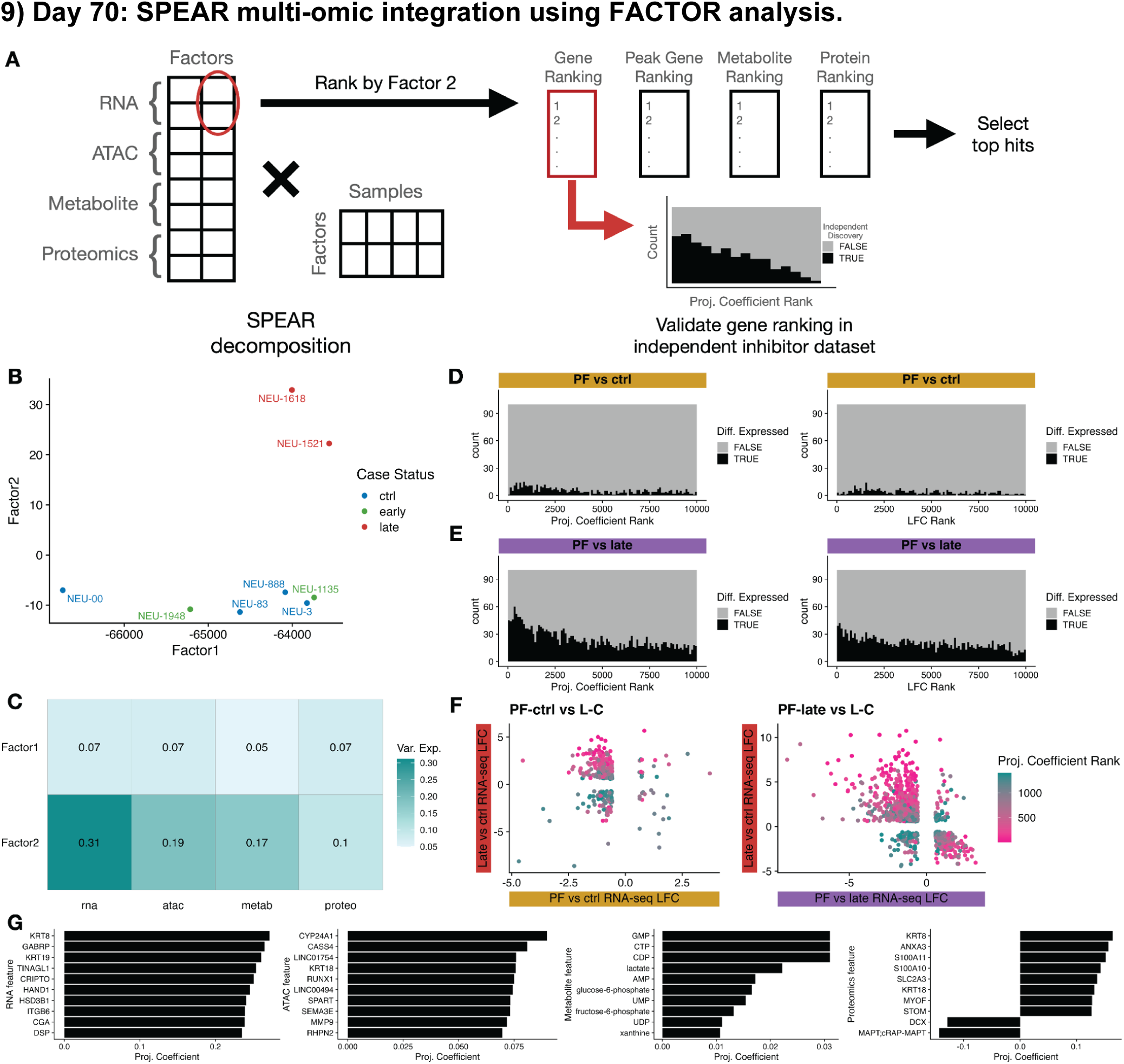
Improved ranking of top dysregulated genes via factor-based integration of ATACseq, RNAseq, Proteomic Mass Spectrometry, & Metabolite Mass Spectrometry from neurons. **a**, Overview of a supervised Bayesian factor analysis on our multi-omic data from early, late, and control neurons for identification of molecular signatures driven by class of KAT6A truncating variants. **b**, Samples plotted in factor space, indicating that Factor 2 provides good separation of late cases from controls and early cases. **c**, Variance explained by the different modalities in each Factor, indicating that the total contribution of the data to Factor 2 is strong. **d-e**, Stacked histograms examining the factor 2 coefficient ranks (left) based on whether or not they were differentially expressed in the KAT6A inhibitor dataset, which provides an orthogonal source of information. The highest ranks (largest coefficients) are enriched for differential expression in the late neuron inhibitor comparison, more so than when comparing the ranks of the log fold changes (right) on just the RNA-seq DEGs (Late vs Ctrl), providing evidence that adding the other modalities is improving the interpretation of the top genes. **f**, Comparison of factor coefficient ranks across 3 neuron RNAseq data sets – with “late vs control” on the Y-axis and either “control: PF9363 vs DMSO” (right) or “late: PF9363 vs DMSO” (left) on the X-axis. Coloration by their factor coefficient ranks show higher ranks match with stronger overlaps between the two comparisons. **g**, The top 10 features in Factor 2 from every modality and their associated coefficients, where a positive coefficient indicates higher measurement along Factor 2 (so higher in late mutation cases).

## Notes

### Competing Interest Statement

The authors have declared no competing interest.

### Author Declarations

This study was approved by the Institutional Review Board (IRB) and the Embryonic Stem Cell Research Oversight (ESCRO) committees at UCLA. All biological samples were collected after informed consent (IRB#11-001087). Patients were recruited under IRB approvals at UCLA, the Sick Kids Hopital, and the University of Bonn; the study conformed to the Declaration of Helsinki, and all participants or their legal guardians gave written informed consent, including explicit consent for publication of identifiable facial photographs.

